# Alveolar Immune Profiling Identifies Distinct Subphenotypes of Acute Respiratory Failure

**DOI:** 10.64898/2026.01.13.26344060

**Authors:** Eric D. Morrell, Ted Liu, Marika Orlov, Leonoor S. Boers, Neha A. Sathe, Chelsea Schmitz, Mallorie Mitchem, Anne Chaize, Lucy Y. Gao, Francis L. Mabrey, Sarah E. Holton, Pavan K. Bhatraju, Akram Khan, Catherine L. Hough, Renee D. Stapleton, Hélène van den Heuvel, JanWillem Duitman, Lieuwe D.J. Bos, Mark M. Wurfel, Carmen Mikacenic

## Abstract

Acute respiratory failure causes millions of deaths worldwide each year, highlighting the need for a better understanding of its pathophysiology and approaches to identify treatment-responsive subphenotypes of patients. Although subphenotypes of acute respiratory failure have been described using peripheral blood biomarkers, it remains unclear whether lung-specific molecular profiles can define biologically and clinically meaningful subphenotypes. In this study, we identified four distinct subphenotypes based on the measurement of twenty-five soluble proteins in alveolar fluid collected from 466 patients with acute respiratory failure. Forty-eight of these participants also underwent immunophenotyping by spectral flow cytometry to characterize immune cell populations. Twenty-eight-day mortality was significantly different across the four subphenotypes. The subphenotype with lowest mortality (12.7%) showed elevated levels of soluble PD-L1, enrichment of T cell-related chemokines, and the highest proportion of memory CD8^+^ T cells. A subphenotype characterized by globally low soluble protein levels and higher proportions of exhausted T cell subsets had a mortality rate of 22.1% despite relatively moderate illness severity. The subphenotype with the highest mortality (29.4%) exhibited increased pyrogenic and neutrophil-associated alveolar mediators. Severity of respiratory failure varied markedly between subphenotypes, while non-pulmonary organ failures were similar across groups. We developed a parsimonious classifier to predict subphenotypes in two additional cohorts totaling 122 patients. The clinical differences between the subphenotypes were similar across cohorts, although associations with mortality were not detected in the smaller validation cohorts. These findings identify novel subphenotypes of acute respiratory failure and support integrating lung-specific molecular measures into future studies to advance precision-medicine approaches.

**One Sentence Summary:** This study identifies novel subphenotypes of acute respiratory failure that exhibit highly distinct immune signatures and associations with clinical outcomes.

## INTRODUCTION

Critical illness resulting from acute respiratory failure is a major cause of morbidity and mortality. Global estimates suggest that hospital mortality amongst mechanically ventilated patients admitted to an intensive care unit (ICU) approach 30% (*1*). The most severe manifestation of acute respiratory failure is acute respiratory distress syndrome (ARDS), which is defined by the clinical criteria of severe hypoxemia accompanied by bilateral pulmonary opacities on chest imaging (*2, 3*). ARDS occurs in roughly 10% of all ICU patients worldwide and is associated with a hospital mortality of 25%-50% (*4*). Patients who are at-risk for ARDS but do not meet full diagnostic criteria (e.g. unilateral pneumonia) still experience mortality of 10-25% (*5–7*). Aside from antimicrobials to treat infections, no therapies directed at the underlying pathophysiologic aspects of acute respiratory failure have demonstrated efficacy.

One of the most important reasons for the lack of successful interventional trials in patients with acute respiratory failure is believed to be the heterogeneity, both clinical and molecular, that is present in patients qualifying for syndromic definitions of disease. For example, only half of patients who meet criteria for ARDS have evidence of diffuse alveolar damage, which is the histologic correlate of ARDS (*8*). Multiple investigators have identified subphenotypes of patients with acute respiratory failure using various types of finite mixture models, plasma biomarker, or transcriptomic clustering approaches (*9–18*). In retrospective observational studies, many of these subphenotypes are associated with differences in mortality and response to therapies.

However, a major limitation of these prior studies has been the lack of accounting for organ-specific (i.e. lung) host responses. It is possible a large degree of heterogeneity in airway and alveolar compartmentalized responses are not being captured within existing critical illness subphenotypes. We and others have shown that peripheral blood molecular signatures are often poor surrogates for lower respiratory tract cellular and molecular biology in patients with acute respiratory failure (*19–26*). In addition, associations with clinical outcomes between lower respiratory tract and peripheral blood molecular signatures may be distinct (*19–21*). Together, these findings highlight a key knowledge gap in our understanding of the relationship between existing acute respiratory failure/ARDS subphenotypes and pulmonary biology. A better understanding of the local factors that drive acute respiratory failure is essential to developing lung-targeted treatments and other precision-based interventional trial strategies.

The primary objective of this study was to identify and characterize molecular subphenotypes of patients with acute respiratory failure using alveolar protein profiles. To achieve this aim, we applied latent profile analysis (LPA) to a panel of 25 soluble proteins reflective of a broad range of immunologic processes we measured in bronchoalveolar lavage fluid (BALF) collected from 466 patients with acute respiratory failure enrolled into the University of Washington Acute Respiratory Failure (UW-ARF) Study. We characterized these novel subphenotypes by testing whether they were associated with clinical factors and outcomes. In a subset of forty-eight subjects, we compared the proportions of alveolar leukocyte subsets between the subphenotypes using spectral flow cytometry. We then tested for the robustness of our results across two independent validation cohorts representing an additional 122 individuals with early (<48 hours) acute respiratory failure.

## RESULTS

### Alveolar Proteins Define Novel Subphenotypes Associated with Distinct Clinical Characteristics

We enrolled 466 mechanically ventilated patients at-risk for or with established ARDS who underwent a bronchoscopy with BAL for suspicion of pneumonia or worsening respiratory status from 2015 – 2023. Individuals with COVID-19 were excluded. The clinical characteristics of individuals enrolled in the UW-ARF Cohort are shown in Table 1. The median age was 52, 26% were female, 32% had shock; the median interval of time between intubation and BAL collection was 6 days, 53.1% of patients had pneumonia, and 55.6% of the cohort met Berlin Criteria for ARDS. Hospital mortality was 25.8%.

**Table 1.**
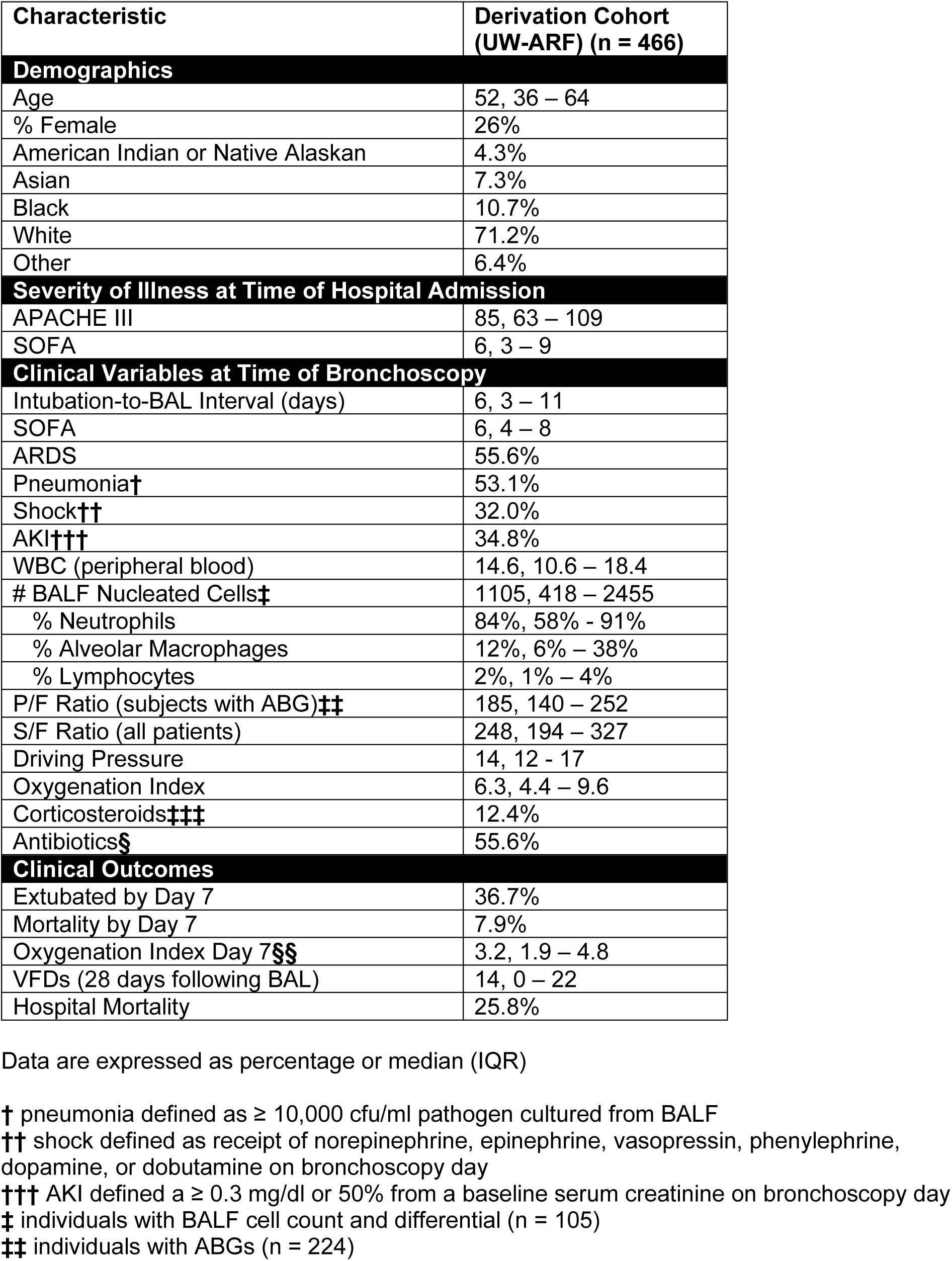
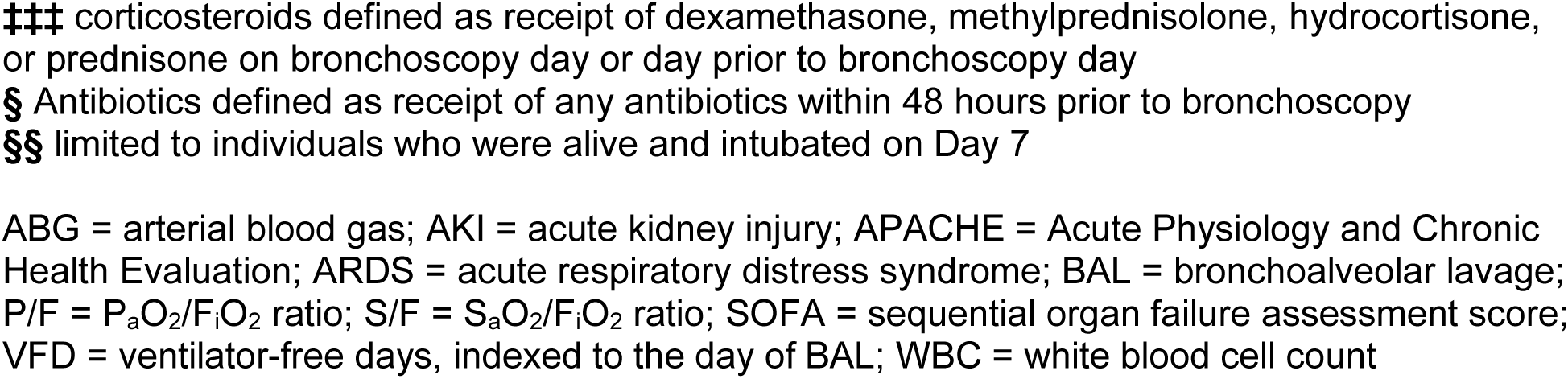
Subject Characteristics in the Derivation Cohort.

We applied LPA to a panel of 25 proteins measured from BAL samples to identify alveolar subphenotypes (A-SPs) (*27*). Alveolar proteins measured below the assay detection limits or not meeting quality control thresholds were excluded from analyses (the original panel included 40 proteins, of which 15 were excluded for not meeting quality control thresholds) (Table S1 and Figure S1). A four subphenotype LPA model provided the best fit (Table S2).

The Vuong-Lo-Mendell-Rubin (VLMR) test indicated that a four-class model was a significant improvement over a two- or three-class model, but a five-class model did not significantly increase how well the classes explained the patterns observed in the data. The value of the BIC plateaued around 4-classes, also suggesting an optimal number of groups. We labeled the four classes A-SP1, A-SP2, A-SP3, and A-SP4.

The alveolar protein profiles were highly distinct between the four A-SPs. Figure 1A displays a heatmap of the standardized values of each of the 25 alveolar proteins between each A-SP. A-SP2 was characterized by high relative expression of chemokines (e.g. CCL13, CCL17, CXCL10), T cell maturation factors (IL-12/IL-23p40), sPD-L1 (checkpoint protein), sCD163 (hemoglobin-haptoglobin receptor), and sRAGE (multi-ligand pattern recognition receptor). A-SP3 had relatively low levels of all 25 proteins, and A-SP4 was distinguished by high pyrogenic (e.g. IL-1α, IL-1β) and neutrophil-associated (e.g. IL-8, S100A8/A9) protein levels. Growth factors IL-7 and VEGF were uniquely elevated in A-SP4. A-SP1 had higher expression of all BAL proteins than A-SP3, but lower chemokines levels compared with A-SP2, and lower inflammatory mediator levels compared with A-SP4.

**Figure 1.**
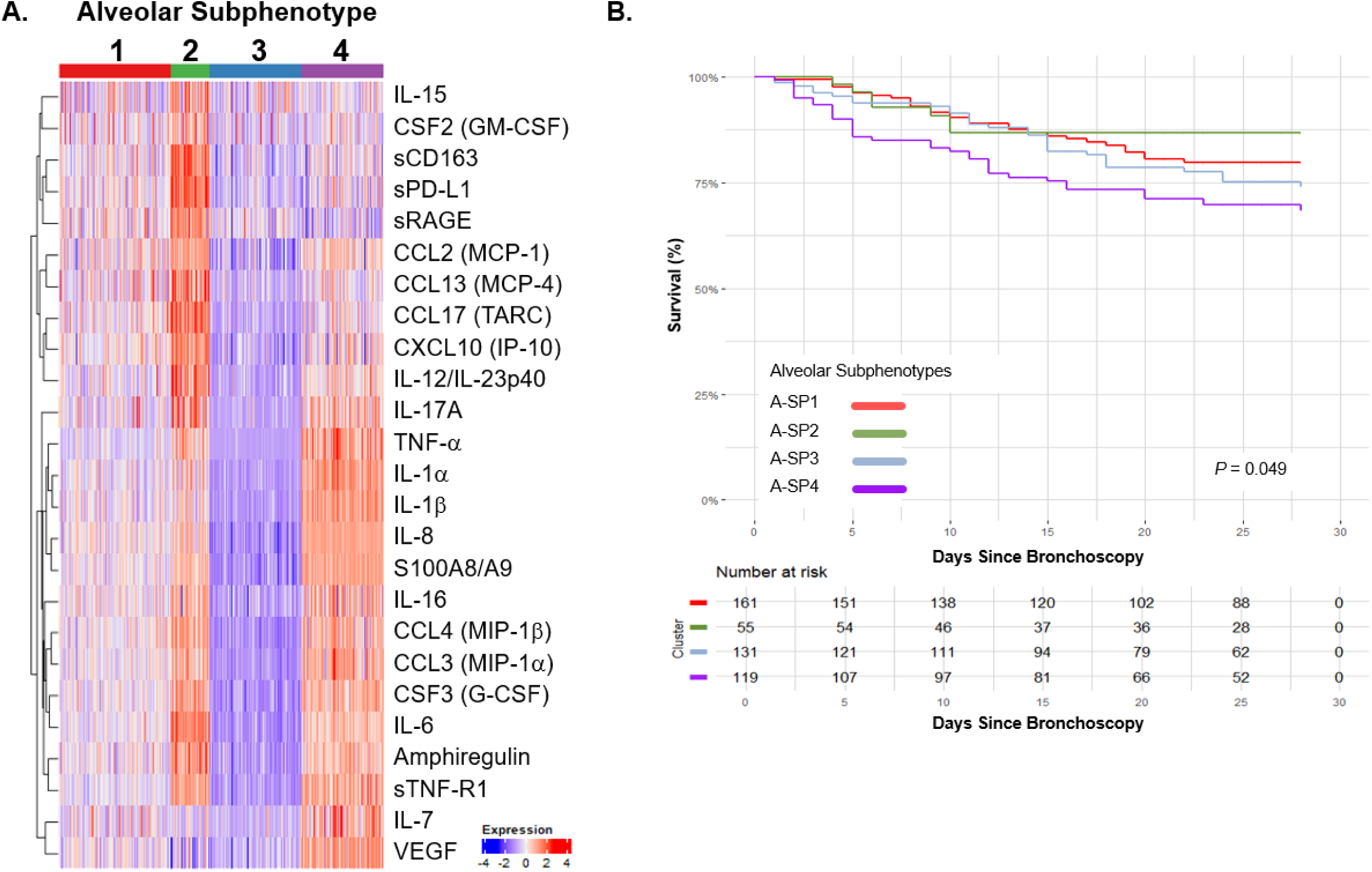
Alveolar Subphenotype Profiles and Associations with Mortality. **A)** Heatmap of standardized values of each protein by alveolar subphenotype (A-SP) in the Derivation Cohort. The variables were standardized by scaling log transformed values to zero with standard deviation = 1. The proteins were sorted on the Y-axis by hierarchical clustering. **B)** Kaplan-Meier survival analysis up to 28 days following bronchoalveolar lavage (BAL) in the Derivation Cohort. *P*-value was generated with a log-rank test. A-SP = alveolar subphenotype

We sought to determine the clinical characteristics that distinguish individuals in each A-SP. Individuals in A-SP2 were significantly younger compared with participants in the other A-SPs (Table 2). The median interval of time between intubation and BAL was also significantly shorter in A-SP2 compared with the other A-SPs (4 days in A-SP2 vs. 6 days in A-SP4, *p* < 0.01). On the day of BAL, the severity of respiratory failure as assessed by ARDS, S/F ratio, and oxygenation index was significantly different between the A-SPs (Figure 2A). The proportion of patients with pneumonia was highest in A-SP4 (80.7%) and lowest in A-SP3 (41.8%).

**Figure 2.**
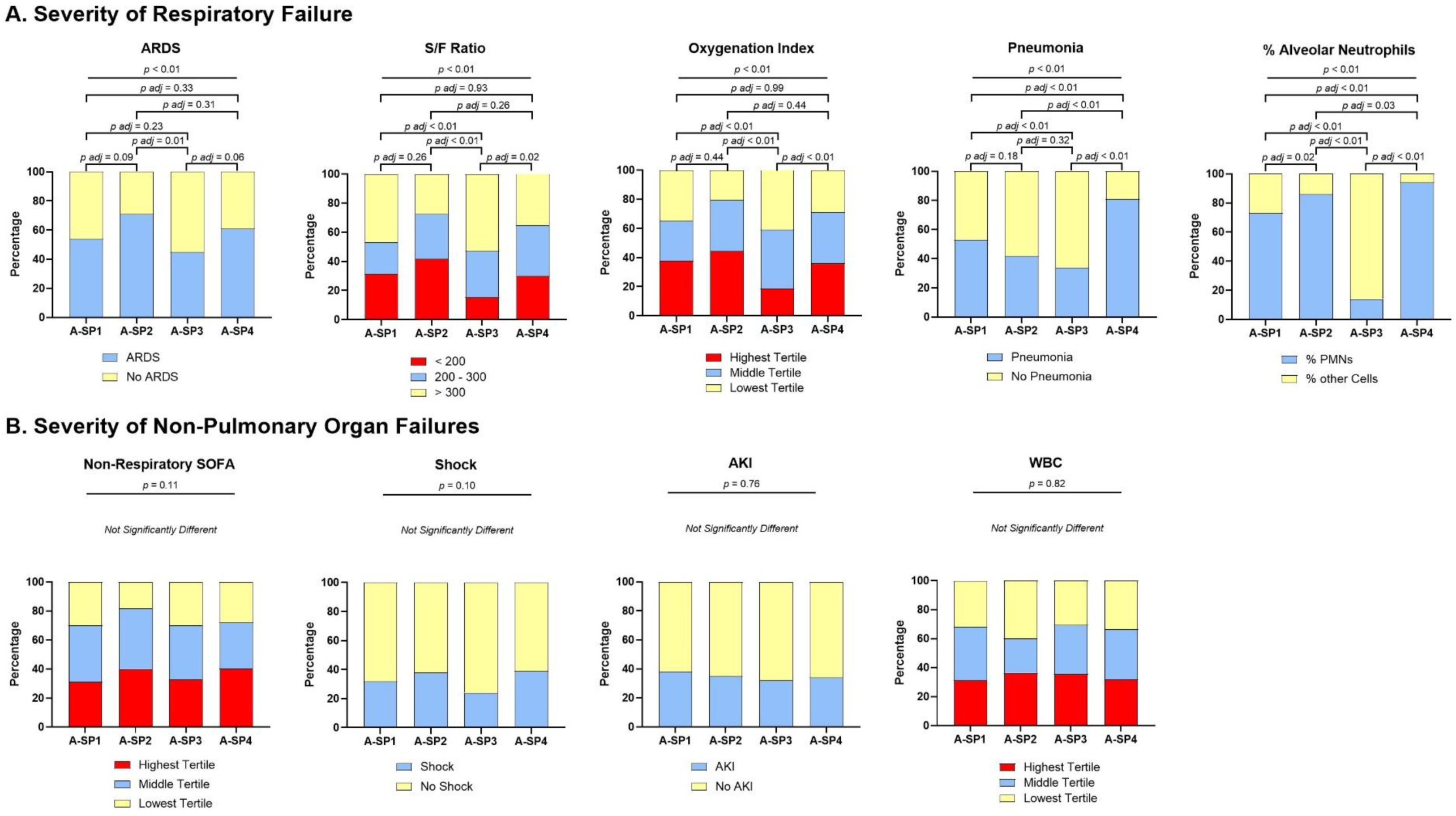
Alveolar Subphenotype Clinical Characteristics. Bar plots of representative pulmonary **(A)** and non-pulmonary **(B)** organ failures. Bar height represents the proportion of each value of the variable. Differences in % ARDS, pneumonia, shock, and AKI were tested with Chi-squared with post-hoc pairwise proportion tests with FDR correction; Differences in % alveolar neutrophils were tested with a Kruskal-Wallis with post-hoc Dunn’s Test with FDR correction; Differences in S/F Ratio, oxygenation index, and Non-Respiratory SOFA were tested with Chi-squared testing differences in the highest (worst) tertile between A-SPs with post-hoc pairwise proportion tests with FDR correction

**Table 2.**
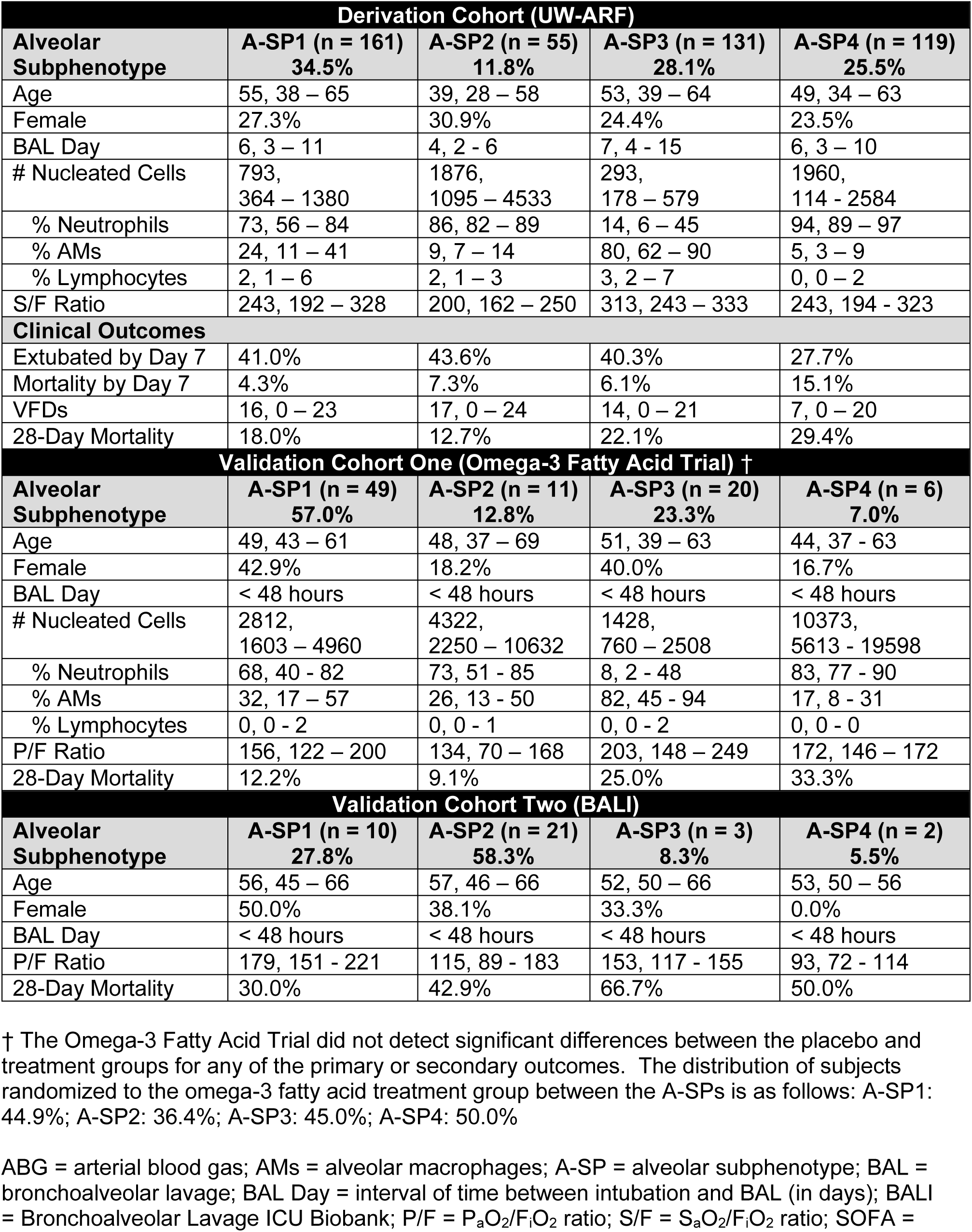
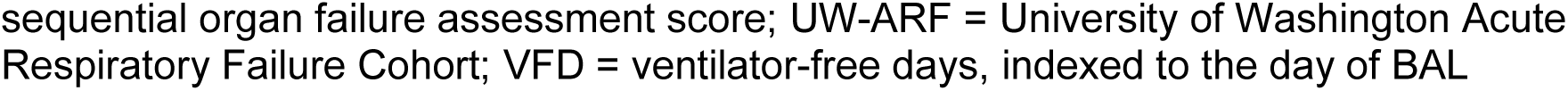
Alveolar Subphenotype Clinical Characteristics and Outcomes.

| **Derivation Cohort (UW-ARF)** |  |  |  |  |
| --- | --- | --- | --- | --- |
| **Alveolar Subphenotype** | **A-SP1 (n = 161)**<br>**34.5%** | **A-SP2 (n = 55)**<br>**11.8%** | **A-SP3 (n = 131)**<br>**28.1%** | **A-SP4 (n = 119)**<br>**25.5%** |
| Age | 55, 38 – 65 | 39, 28 – 58 | 53, 39 – 64 | 49, 34 – 63 |
| Female | 27.3% | 30.9% | 24.4% | 23.5% |
| BAL Day | 6, 3 – 11 | 4, 2 - 6 | 7, 4 - 15 | 6, 3 – 10 |
| # Nucleated Cells | 793,<br>364 – 1380 | 1876,<br>1095 – 4533 | 293,<br>178 – 579 | 1960,<br>114 - 2584 |
| % Neutrophils | 73, 56 – 84 | 86, 82 – 89 | 14, 6 – 45 | 94, 89 – 97 |
| % AMs | 24, 11 – 41 | 9, 7 – 14 | 80, 62 – 90 | 5, 3 – 9 |
| % Lymphocytes | 2, 1 – 6 | 2, 1 – 3 | 3, 2 – 7 | 0, 0 – 2 |
| S/F Ratio | 243, 192 – 328 | 200, 162 – 250 | 313, 243 – 333 | 243, 194 - 323 |
| **Clinical Outcomes** |  |  |  |  |
| Extubated by Day 7 | 41.0% | 43.6% | 40.3% | 27.7% |
| Mortality by Day 7 | 4.3% | 7.3% | 6.1% | 15.1% |
| VFDs | 16, 0 – 23 | 17, 0 – 24 | 14, 0 – 21 | 7, 0 – 20 |
| 28-Day Mortality | 18.0% | 12.7% | 22.1% | 29.4% |
| **Validation Cohort One (Omega-3 Fatty Acid Trial) †** |  |  |  |  |
| **Alveolar Subphenotype** | **A-SP1 (n = 49)**<br>**57.0%** | **A-SP2 (n = 11)**<br>**12.8%** | **A-SP3 (n = 20)**<br>**23.3%** | **A-SP4 (n = 6)**<br>**7.0%** |
| Age | 49, 43 – 61 | 48, 37 – 69 | 51, 39 – 63 | 44, 37 - 63 |
| Female | 42.9% | 18.2% | 40.0% | 16.7% |
| BAL Day | < 48 hours | < 48 hours | < 48 hours | < 48 hours |
| # Nucleated Cells | 2812,<br>1603 – 4960 | 4322,<br>2250 – 10632 | 1428,<br>760 – 2508 | 10373,<br>5613 - 19598 |
| % Neutrophils | 68, 40 - 82 | 73, 51 - 85 | 8, 2 - 48 | 83, 77 - 90 |
| % AMs | 32, 17 – 57 | 26, 13 - 50 | 82, 45 - 94 | 17, 8 - 31 |
| % Lymphocytes | 0, 0 - 2 | 0, 0 - 1 | 0, 0 - 2 | 0, 0 - 0 |
| P/F Ratio | 156, 122 – 200 | 134, 70 – 168 | 203, 148 – 249 | 172, 146 – 172 |
| 28-Day Mortality | 12.2% | 9.1% | 25.0% | 33.3% |
| **Validation Cohort Two (BALI)** |  |  |  |  |
| **Alveolar Subphenotype** | **A-SP1 (n = 10)**<br>**27.8%** | **A-SP2 (n = 21)**<br>**58.3%** | **A-SP3 (n = 3)**<br>**8.3%** | **A-SP4 (n = 2)**<br>**5.5%** |
| Age | 56, 45 – 66 | 57, 46 – 66 | 52, 50 – 66 | 53, 50 – 56 |
| Female | 50.0% | 38.1% | 33.3% | 0.0% |
| BAL Day | < 48 hours | < 48 hours | < 48 hours | < 48 hours |
| P/F Ratio | 179, 151 - 221 | 115, 89 - 183 | 153, 117 - 155 | 93, 72 - 114 |
| 28-Day Mortality | 30.0% | 42.9% | 66.7% | 50.0% |

† The Omega-3 Fatty Acid Trial did not detect significant differences between the placebo and treatment groups for any of the primary or secondary outcomes. The distribution of subjects randomized to the omega-3 fatty acid treatment group between the A-SPs is as follows: A-SP1: 44.9%; A-SP2: 36.4%; A-SP3: 45.0%; A-SP4: 50.0%
ABG = arterial blood gas; AMs = alveolar macrophages; A-SP = alveolar subphenotype; BAL = bronchoalveolar lavage; BAL Day = interval of time between intubation and BAL (in days); BALI = Bronchoalveolar Lavage ICU Biobank; P/F = $P_aO_2/F_iO_2$ ratio; S/F = $S_aO_2/F_iO_2$ ratio; SOFA =
sequential organ failure assessment score; UW-ARF = University of Washington Acute Respiratory Failure Cohort; VFD = ventilator-free days, indexed to the day of BAL

We analyzed the clinical BALF cell count and differentials to see if there were immune cell differences between the A-SPs that corresponded with the soluble protein profiles. The % BALF neutrophils was significantly higher in A-SP4 compared with the three other A-SPs, including A-SP2 (*p* < 0.01) (Figure 2A). These differences in the number and proportion of neutrophils between the A-SPs mirrored the relative levels of neutrophil-related mediators like IL-8 and S100A8/A9 reflected in the soluble protein profiles (Figure 1A). The % BALF lymphocytes was highest in A-SP3, and median % BAL lymphocytes was significant higher in A-SP2 (2.0%, 1.0 – 3.3%) compared with A-SP4 (0.0, 0.0 – 2.0) (*p* < 0.01) (Table 2).

In contrast to the differences in severity of respiratory failure and alveolar neutrophilia we observed between the A-SPs, we did not detect significant differences in the severity of non-pulmonary organ failures such as shock, acute kidney injury (AKI), and peripheral white blood cell count between A-SPs (Figure 2B). These findings support the notion that A-SPs reflect lung-specific pathophysiology rather than systemic illness severity.

### Alveolar Subphenotypes have Distinct Associations with Mortality

We proceeded to investigate whether A-SPs were associated with mortality in patients with acute respiratory failure. The proportion of patients alive and extubated by day 7 following the BAL was highest in A-SP2 (43.6%) and lowest in A-SP4 (27.7%) (Table 2). The number of days alive and free of mechanical ventilation (VFD) in the 28 days following the BAL was highest in A-SP2 (VFDs = 17) and lowest in A-SP4 (VFDs = 7) (*p* = 0.03) (Table 2). Kaplan-Meier analysis showed an association between A-SPs and mortality within 28 days following the BAL (log-rank *p* = 0.049) (Figure 1B). Individuals in A-SP4 had the lowest, whereas A-SP2 had the highest, respective survival in the 28 days following BAL amongst the A-SPs. These findings suggest that two extremes of an alveolar immune response – unopposed innate immune activation (A-SP4: IL-1β, IL-8, S100A8/A9, monocytes, neutrophils) or a highly dampened response (A-SP3) – are most strongly associated with poor outcomes in acute respiratory failure. In contrast, a host response reflective of moderate innate immune activation balanced with T cell and regulatory mediators (A-SP2) is associated with improved survival.

We performed multiple sensitivity analyses to better understand the differences in mortality between the A-SPs. Subjects in A-SP2 had the lowest mortality in early (0 – 6 days), middle (7 – 14 days), or late (> 14 days) occurring BALs (Table S3). In contrast, subjects in A-SP4 had the highest mortality regardless of when their BAL occurred relative to intubation. A-SP4 also had the highest mortality rate regardless of age. A-SP2 had the lowest mortality rate in younger individuals, but this difference was attenuated in older participants. Individuals in A-SP2 had the lowest, whereas A-SP4 had the highest, mortality rates irrespective of pneumonia status. Interestingly, A-SP2 had the lowest mortality in subjects with ARDS, but the highest mortality in non-ARDS subjects (although this was limited to only 15 individuals). Exposure to corticosteroids or antibiotics prior to bronchoscopy did not dramatically change the distribution of mortality between the A-SPs. Finally, we developed a relative risk regression model testing whether A-SPs were associated with mortality adjusting for age, sex, pneumonia, and ARDS (Figure S2). A-SP4 was associated with a 2.20 relative risk for death compared with A-SP2 in this model (*p* < 0.05). Including intubation-to-BAL interval into this full model attenuated the association to the threshold of statistical significance (*p* = 0.05). Together, these sensitivity analyses suggest the mortality differences between the A-SPs, especially A-SP2 and A-SP4, are not completely driven by some of the most common clinical characteristics used to classify patients with acute respiratory failure such as pneumonia and ARDS.

### Lower Respiratory Immune Cell Phenotypes are Distinct Between Alveolar Subphenotypes

To better understand the alveolar cellular immune responses underlying the A-SPs, we analyzed spectral flow cytometry data of BALF from a subset of 48 participants included in the UW-ARF Study. Table S4 displays the clinical characteristics of subjects in this subset of the UW-ARF Cohort. The selection of these 48 samples was based on research staff availability at the time of the bronchoscopy to process the BALF sample for cell cryopreservation. Our cell-surface protein panel was designed to capture alveolar myeloid cell subsets previously identified in patients with acute respiratory failure (CD14^+^CD206^-^ monocytes, CD206^+^CD71^HI^ macrophages, CD206^+^CD71^LO^CD163^HI^ macrophages) (*21, 22, 28, 29*), CD4^+^CD25^HI^CD127^LO^ regulatory T cells (Tregs) (*30*), T cell subsets on the “exhaustion” spectrum (PD-1^HI^) (*21, 31*), innate-like T cell subsets (γδ T cells, MAIT cells) (*32, 33*), and cell subsets whose soluble ligands were included in A-SP soluble protein profiles (CCR2:CCL2; CXCR3:CXCL10; CCR4:CCL17; PD-1:sPD-L1; CD127:IL-7) (Table S5 and Figure S3).

The proportions of CD14^+^CD206^-^ alveolar monocytes (as well as the CCR2^HI^ monocytes) were higher in A-SP2 and A-SP4 compared with A-SP1 and A-SP3 (Figure 3A). Inversely, the proportion of the CD206^+^CD71^HI^CD163^HI^ mature macrophage subset was highest in A-SP1 and A-SP3. These findings correspond with the higher CCL2 BALF levels in A-SP2 and A-SP4 (Spearman’s *r* = 0.35 for BALF CCL2 levels and % CD14^+^CD206^-^ alveolar monocytes in the entire cohort (n = 47)). Notably, the proportion of γδ T cells was significantly higher in A-SP2 compared with A-SP4 (Figure 3B). γδ T cells, a minor T cell lineage that are enriched in peripheral tissues, have previously been shown to protect against either pneumococcal or pseudomonas-induced lung injury in mice through improved bacterial clearance (*32, 33*). There were no differences in the proportion of MAIT cells across A-SPs.

**Figure 3.**
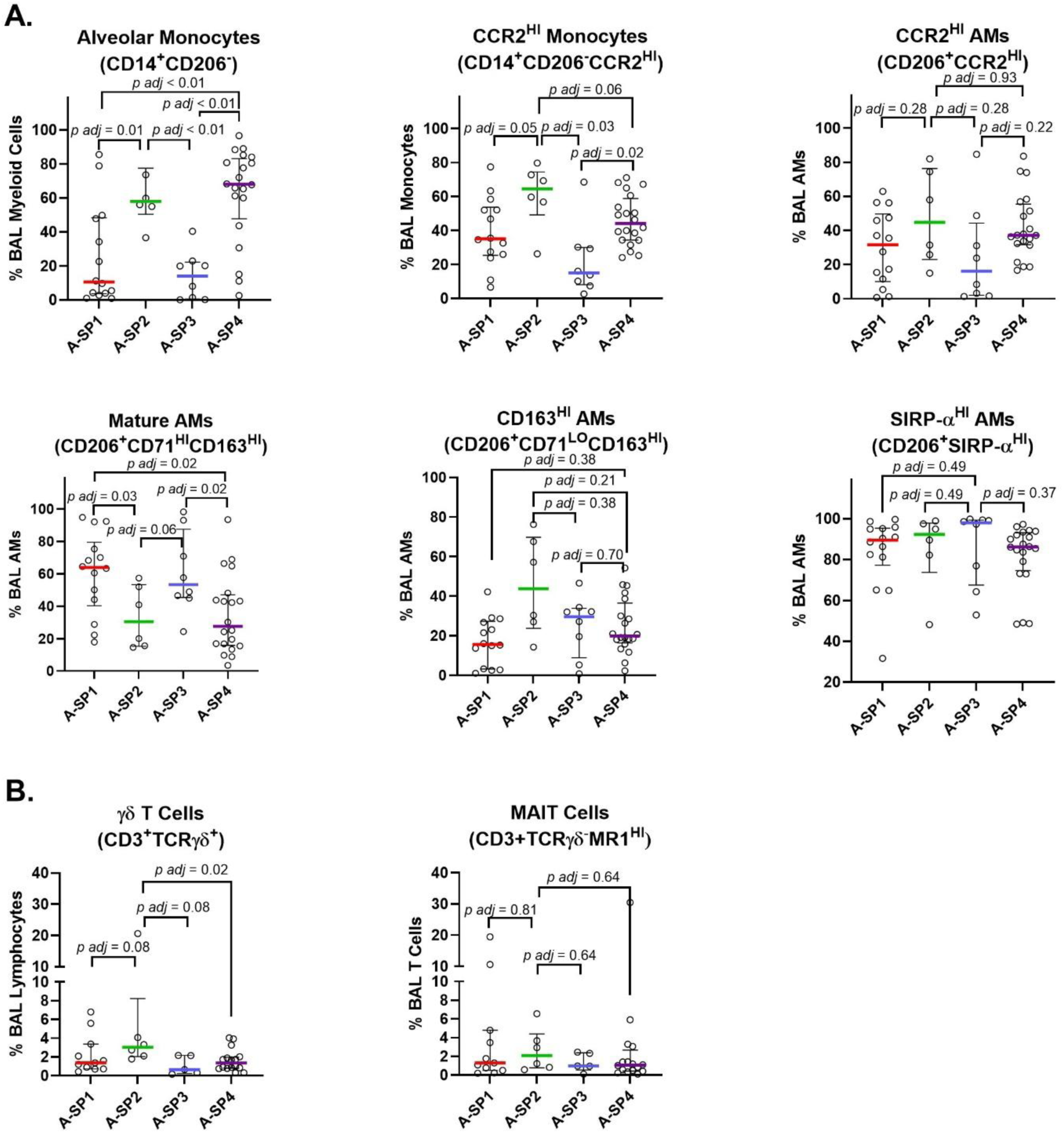
Myeloid and Innate-Like T Cell Subset Proportions Between Alveolar Subphenotypes. Panels show the percentage of alveolar **A)** myeloid and **B)** innate-like T cell subsets as a proportion of their respective parent populations. Shown are individual percentages (dots), medians (lines), and interquartile ranges (error bars). *P*-values were calculated with Mann-Whitney rank sum tests and adjusted for the number of tests performed per subset by the Benjamini-Hochberg (BH) approach (*p adj*). AM = alveolar macrophage; A-SP = alveolar subphenotype; BAL = bronchoalveolar lavage; MAIT = mucosa-associated invariant T cells

The median CD4^+^/CD8^+^ T cell ratio was highest in A-SP2, and was significantly higher in A-SP2 (ratio = 3.8) vs. A-SP4 (ratio = 1.5) (*p* = 0.02). The cellular profile of A-SP3 (which was characterized by the lowest relative expression of almost all soluble proteins measured) was marked by relatively high proportions of CXCR3 expressing CD4^+^ and CD8^+^ T cells on the T_H_1 spectrum (Figure 4A and 4B) (*31*). CCR6^+^ T cells were also more abundant in A-SP1 and A-SP3 compared with A-SP2 and A-SP4. We did not detect a significant difference in the proportion of CD4^+^CD25^HI^CD127^LO^ Tregs between any of the A-SPs, however the proportion of CD8^+^CD127^HI^ T cells was significantly higher in A-SP2 compared with the other A-SPs (*34, 35*). In summary, the cytometric profiles between the different A-SPs suggest the following: A-SP2: high proportions of active (γδ) and memory (CD8^+^CD127^HI^) T cell subsets with low proportions of exhausted or effector T cell subsets (CCR6^HI^ and PD-1^HI^); A-SP3: high proportions of T_H_1 CXCR3^HI^ T cells; and A-SP4: high proportions of monocytes and neutrophils.

**Figure 4.**
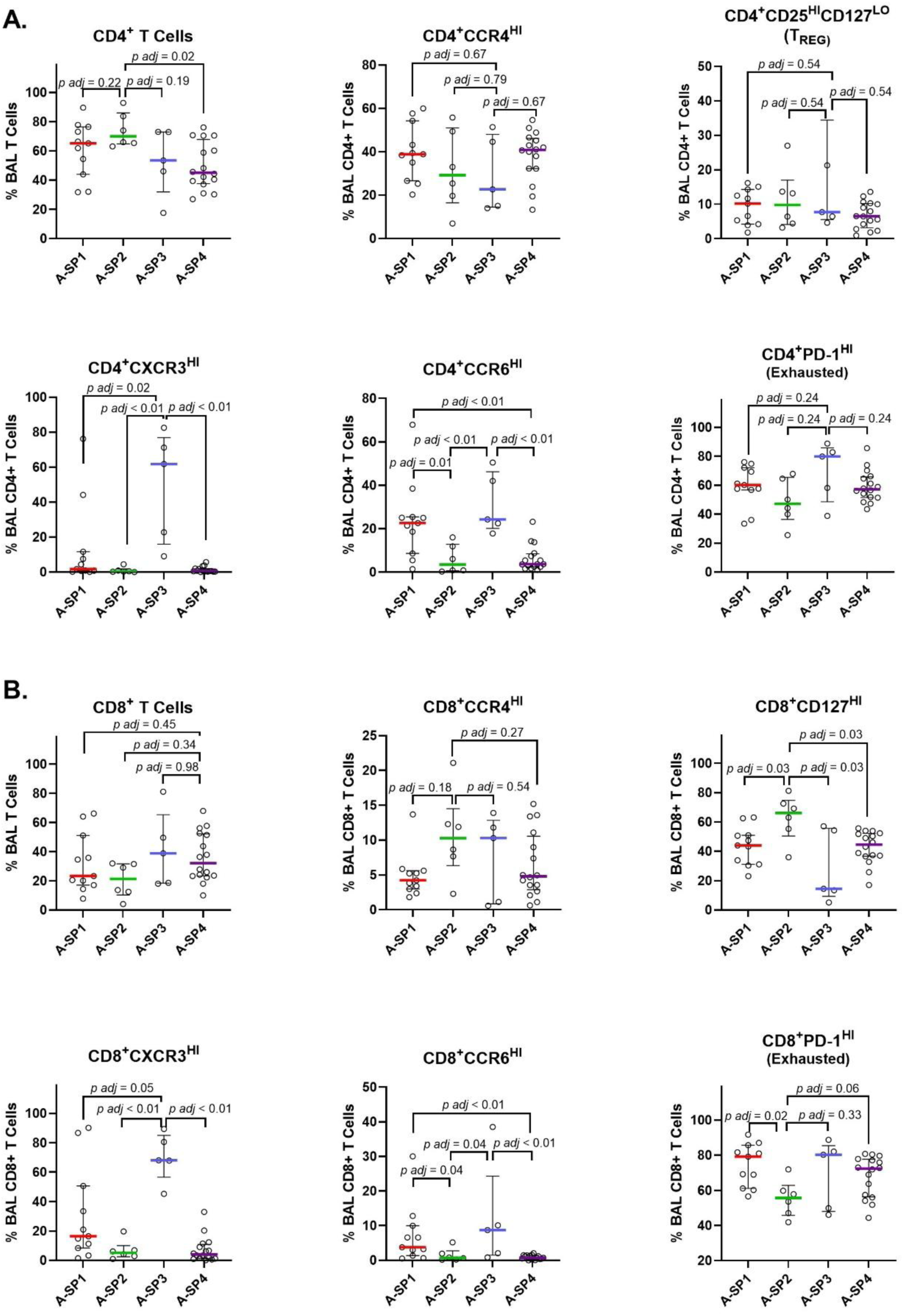
T Cell Subset Proportions Between Alveolar Subphenotypes. Panels show the percentage of alveolar **A)** CD4^+^ and **B)** CD8^+^ T cell subsets as a proportion of their respective parent populations. Shown are individual percentages (dots), medians (lines), and interquartile ranges (error bars). *P*-values were calculated with Mann-Whitney rank sum tests and adjusted for the number of tests performed per subset by the Benjamini-Hochberg (BH) approach (*p adj*). A-SP = alveolar subphenotype; BAL = bronchoalveolar lavage

### Validation of Alveolar Subphenotypes in External Cohorts

We next sought to determine whether we could identify A-SPs in two external cohorts of patients with early (< 48 hours from the onset of mechanical ventilation) acute respiratory failure given that the median time from intubation to bronchoscopy was 6 days in UW-ARF (Table 1). Validation Cohort One consisted of patients (n = 86) enrolled in the Phase II randomized controlled trial of omega-3 fatty acids vs. placebo for treatment of ARDS (*36*). Enrolled subjects from five North American Centers underwent a research bronchoscopy with BAL prior to study drug administration. Validation Cohort Two consisted of patients (n = 36) enrolled in the BAL ICU Biobank (BALI) Study conducted at two hospitals that are part of the Amsterdam University Medical Center (Amsterdam UMC). These subjects underwent clinical bronchoscopy and BAL to help determine the etiology of acute respiratory failure. The clinical characteristics of subjects in Validation Cohorts One and Two are shown in Table S6. Compared with the UW-ARF (Derivation Cohort), Validation Cohort One had a lower mortality rate (16.3%) whereas Validation Cohort Two had a higher mortality rate (41.7%).

We used a gradient-boosted machine algorithm (xGBoost) to develop a classifier model from Derivation Cohort data and interpreted the estimates using Shapley additive explanation (SHAP) values (Figure S4). The SHAP summary plot showed that IL-6, sPD-L1, S100A8/A9 (calprotectin), and IL-8 were the top contributors to the likelihood of assignment to A-SP1, A-SP2, A-SP3, and A-SP4, respectively, in the Derivation Cohort. We then developed a parsimonious model using a smaller set of BALF measurements made in the validation cohorts. Validation Cohort One had existing BALF measures of IL-6, sPD-L1, IL-8, and CSF3 (G-CSF) (the 2^nd^ highest SHAP value for A-SP3). Validation Cohort Two had existing BALF measures of IL-6, sPD-L1, CSF3 (G-CSF), and VEGF (the 2^nd^ highest SHAP value for A-SP4). Each of these respective 4-biomarker models had good performance for predicting correct A-SP in the Derivation Cohort (each with ROCs values > 0.96 - Figure S5), therefore we carried forward these models to classify A-SP in each of the validation cohorts.

We sought to determine whether the associations between the A-SPs and clinical characteristics we identified in the Derivation Cohort were generalizable to cohorts exclusively comprised of patients with early acute respiratory failure. The standardized alveolar protein profiles in the two validation cohorts are shown in Figures 5A and 5B (Figure S6 displays the non-standardized concentrations of the proteins between each A-SP amongst the cohorts). The proportion of patients in A-SP2 and A-SP3 was similar between the Derivation Cohort and Validation Cohort One, while the percentage of patients in A-SP4 was 3.5 times higher in the Derivation Cohort (Table 2). Almost 60% of patients in Validation Cohort Two were in A-SP2, which was much higher than the other two cohorts.

**Figure 5.**
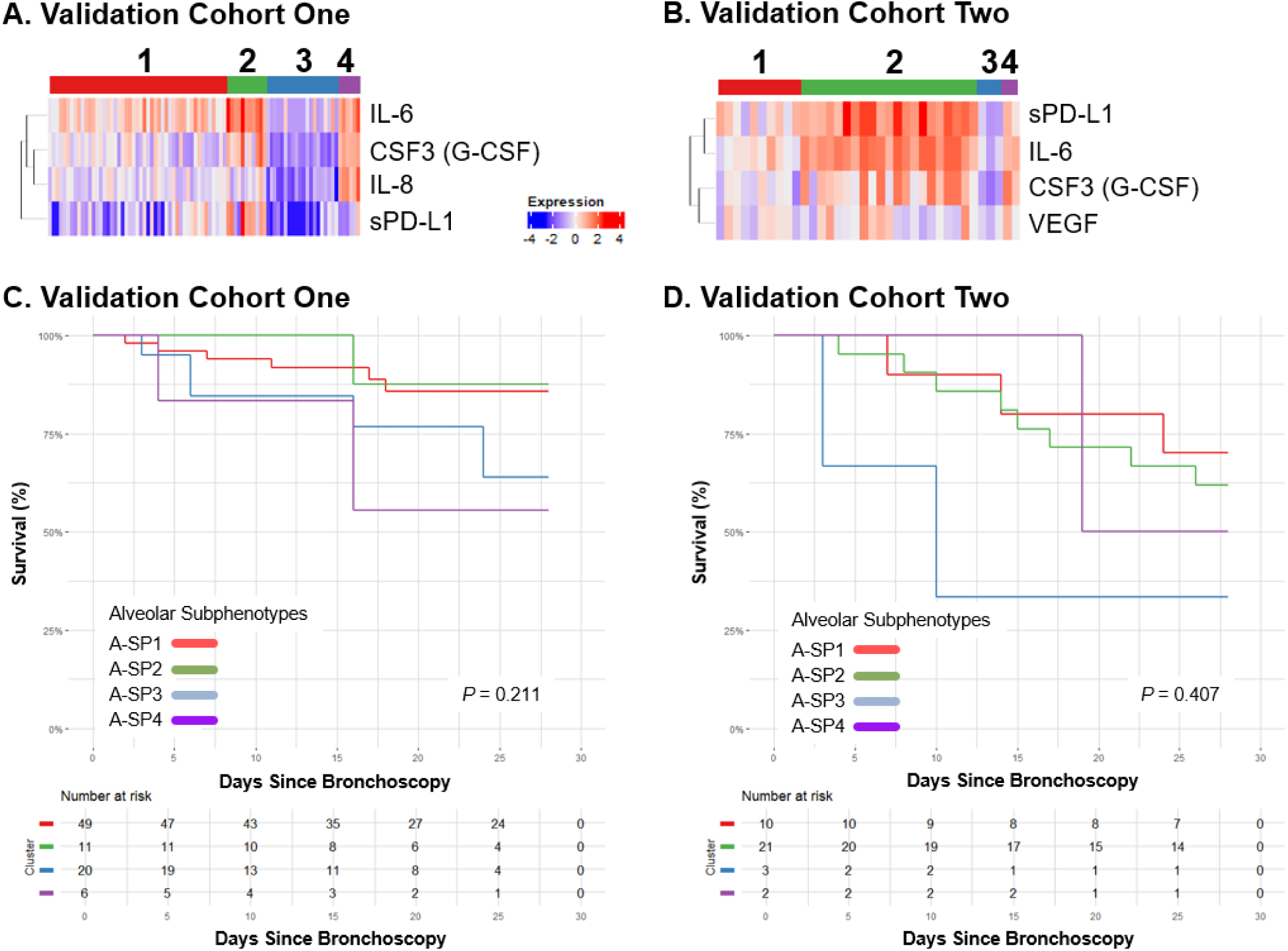
Alveolar Subphenotype Profiles and Associations with Mortality in Validation Cohorts A and. **B)** Heatmaps of standardized values of each protein by alveolar subphenotype (A-SP) in the two Validation Cohorts. The variables were standardized by scaling all log transformed values to zero with standard deviation = 1. The proteins were sorted on the Y-axis by hierarchical clustering. **C and D)** Kaplan-Meier survival analysis to 28 days following bronchoalveolar lavage (BAL) in the Validation Cohorts. *P*-values generated with a log-rank test. A-SP = alveolar subphenotype

The distribution of % BALF neutrophils, number of nucleated cells, degree of hypoxemia, and mortality amongst the four A-SPs in the two validation cohorts were similar to the distributions observed in the Derivation Cohort (Table 2). Specifically, mortality was numerically lowest in A-SP1 and A-SP2 and highest in A-SP3 and A-SP4 in both validation cohorts. Kaplan-Meier analyses did not demonstrate associations between A-SPs and mortality in either of the validation cohorts that met our prespecified threshold for statistical significance (Validation Cohort One: log-rank *p* = 0.21; Validation Cohort Two: log-rank *p* = 0.41).

### Alveolar Subphenotypes are Distinct from Plasma-Derived Critical Illness Subphenotypes

We wanted to determine the degree of overlap between the most widely cited peripheral blood-based ARDS subphenotype classification (“hyper-” and “hypoinflammatory” ARDS) (*18*) and the A-SPs we derived from lower respiratory tract measurements. All subjects in Validation Cohort One had ARDS and paired plasma samples, therefore we were able to use data from this cohort to address this question. There were significant differences in the proportion of hyperinflammatory ARDS patients between the A-SPs (Figure S7). A-SP2 and A-SP4 had the highest proportion of hyperinflammatory ARDS patients (73% of A-SP2 subjects were hyperinflammatory; 50% of A-SP4 subjects were hyperinflammatory).

### Patients Change Alveolar Subphenotype Assignment Over the Course of a Hospitalization

We explored the degree that individuals within A-SPs changed subphenotypes over the course of a hospitalization, and whether these changes were associated with mortality. Twenty-five subjects in our Derivation Cohort had a repeat bronchoscopy performed during their hospital course and two individuals had a third bronchoscopy. The median interval of time between the first and second bronchoscopies was 14 days (Figure 6). The majority of patients changed A-SP over their hospital course. Most individuals (86%) who were in A-SP4 on the second bronchoscopy had pneumonia at that timepoint. We did not detect any clear trends between A-SP trajectory and hospital mortality, although both subjects who transitioned from A-SP1 to A-SP4 died and the two individuals who had a third bronchoscopy both eventually transitioned into A-SP3.

**Figure 6.**
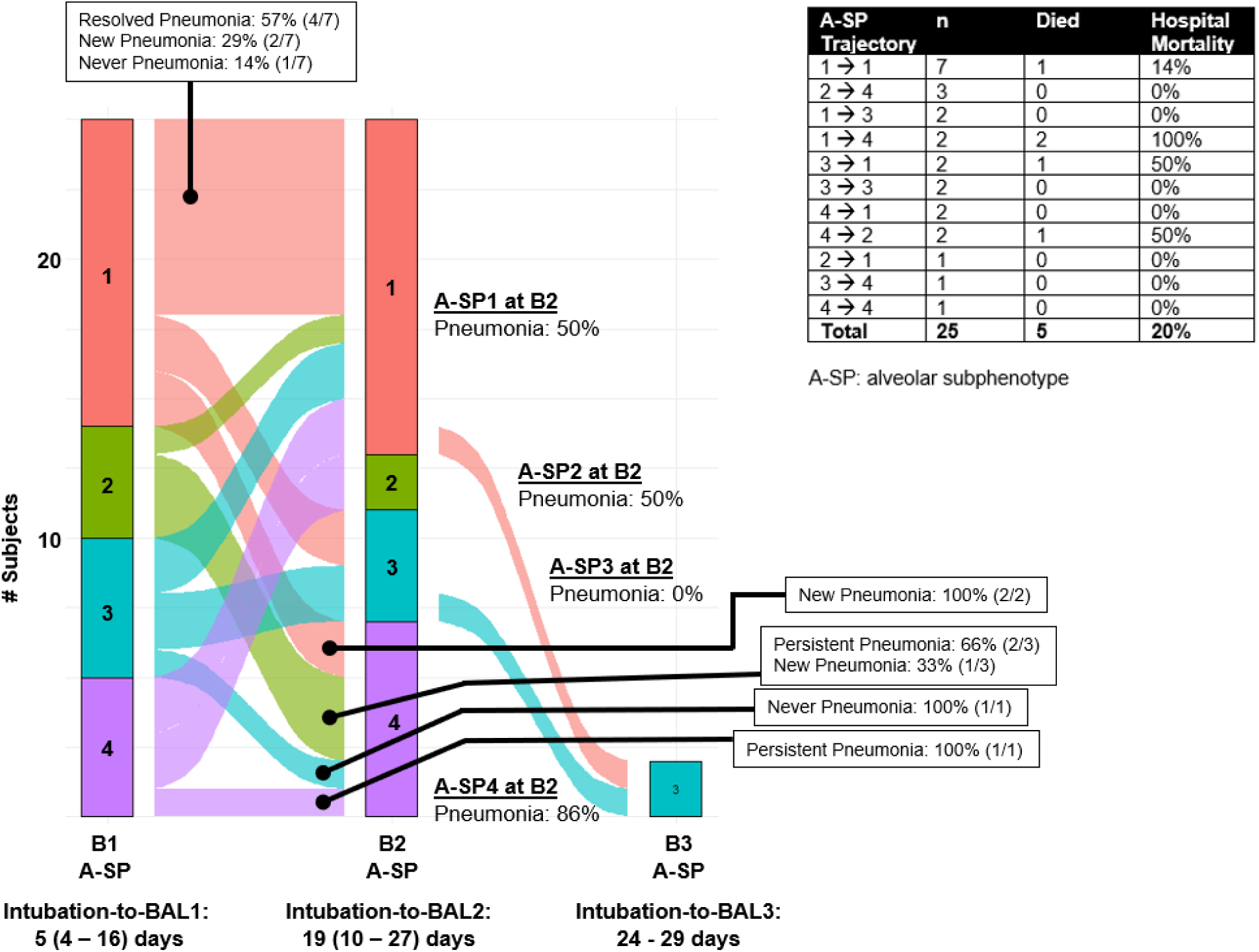
Alveolar Subphenotype Trajectories Over Time. Alluvial diagram displays the number of individuals assigned to each A-SP at bronchoscopy 1 (B1), bronchoscopy 2 (B2), or bronchoscopy 3 (B3) (X-axis). Y-axis displays the number of subjects in A-SP1 (red), A-SP2 (green), A-SP3 (teal), and A-SP4 (purple).

## DISCUSSION

Our finding that subphenotypes derived from alveolar soluble protein measurements in patients with acute respiratory failure have distinct immune profiles and clinical outcomes significantly expands current conceptual models of critical illness syndromic classification. Although prior studies have shown that individual alveolar mediators (such as IL-8 and Tregs) play important roles in acute respiratory failure, our analysis is the first to derive subphenotypes using alveolar immune profiles from multiple large, well-characterized, clinical cohorts. We demonstrate that mediators measured from the point of injury – the lower respiratory tract – are a key source of heterogeneity in acute respiratory failure, and that this heterogeneity might help identify patient subgroups with unique biology and potentially distinct therapeutic responses.

Our findings support the idea that an unbalanced immunologic response in the alveolar space during acute respiratory failure may contribute to early mortality. Consistent with this, we found that A-SP3 and A-SP4 – subphenotypes with nearly opposite immune profiles – were associated with the highest mortality across all three cohorts. A-SP4 was characterized by neutrophilic and monocytic alveolitis and by elevated levels of innate immune cytokines (e.g. IL-1β, TNF-α) and chemokines (e.g. IL-8) that have previously been associated with ARDS- and severe pneumonia-related outcomes (*28, 37–40*). In contrast, A-SP3 displayed uniformly low BALF concentrations across almost all 25 proteins we measured. To our knowledge, this type of blunted alveolar immune profile has not been described in a large cohort of critically ill patients with acute respiratory failure, although immunoparalysis has been described in peripheral blood leukocytes and splenic tissue from patients with severe sepsis (*41–44*). A-SP3 may represent a localized “immunologically exhausted” phenotype that is susceptible to poor outcomes. Future studies are needed to determine whether patients with this alveolar immune profile are more susceptible to viral reactivations (e.g. HSV, CMV) or other opportunistic infections compared with individuals in other A-SPs.

In contrast to an unbalanced alveolar immunologic response being associated with poor outcomes in acute respiratory failure, we found that a T cell recruitment signature balanced with moderate innate immune activation and checkpoint protein expression may be linked with early survival. Subjects in A-SP2 had the lowest mortality rate in both the Derivation Cohort and Validation Cohort One, despite having the highest rates of ARDS and the most severe hypoxemia. Interestingly, subjects in A-SP2 exhibited higher levels of many innate immune cytokines and chemokines typically considered injurious in ARDS (e.g. IL-1β and IL-8) compared with subjects in A-SP1 and A-SP3. Yet compared with A-SP4, subjects in A-SP2 had a more balanced profile with elevated levels of T cell chemokines (e.g. CCL13, CCL17, CXCL10) and growth factors (e.g. IL-12/IL-23p40) that were higher in parallel with moderate elevations in innate immune mediators. Very few studies have measured alveolar T cell chemokine levels in patients with acute respiratory failure or ARDS, although animal models have suggested a key role for various T cell subsets such as Tregs in resolution of inflammation, efferocytosis, and epithelial repair (*30, 45–48*). We also observed that A-SP2 had the highest BALF levels of the checkpoint protein sPD-L1 relative to the other A-SPs (Figure S6). These findings are consistent with prior work demonstrating associations between PD-L1 activity in the lung (soluble protein, cell-surface, and alveolar macrophage expression) and improved outcomes in patients with ARDS (*21, 29*). In summary, our analyses suggest there is a wide range of alveolar immunologic profiles in patients with acute respiratory failure and identifies multiple potential therapeutic targets.

Our corresponding cellular immunophenotyping analyses help contextualize the soluble protein and clinical associations within alveolar biology. Supporting the hypothesis that A-SP3 may represent an immunologically “exhausted” phenotype, we found that surface expression of CXCR3 was highly upregulated on CD4^+^ and CD8^+^ T cells in A-SP3 patients (Figure 4). Consensus guidelines identify CXCR3 as a marker for T_H_1 cells but could also suggest precursor-exhausted T cells (T_PEX_) along with other proteins such as PD-1 (*31*). Although not statistically significant, patients within A-SP3 also had the highest numeric proportions of PD-1 positive CD4^+^ and CD8^+^ T cells as well as SIRP-α^HI^ (“don’t eat me”) alveolar macrophages (Figures 3 and 4). The proportions of alveolar neutrophils and monocytes was highest in A-SP4 – the subphenotype with the worst outcomes – which is consistent with many COVID-19 (*49–52*) and non-COVID-19 (*28, 37*) studies associating this cellular profile with poor outcomes in acute respiratory failure. This pattern may again reflect an uncoordinated and non-specific host attempt at pathogen clearance, given the lack of T cell chemokine and adaptive immune memory cells in A-SP4 compared with A-SP2.

Contrasting the cellular exhaustion signature observed in A-SP3 and the hyperinflammatory signature in A-SP4, our analysis found that subjects in A-SP2 had a significantly higher proportion of CD8^+^CD127^HI^ T cells compared with the other A-SPs (Figure 4B). Although our study excluded individuals with SARS-CoV-2 infection, these findings are consistent with studies in COVID-19 pneumonia and suggest that alveolar T cell enrichment may enhance pathogen clearance or protect against bacterial pneumonia in patients at high risk within A-SP2. Both Markov et al. and Szabo et al. found that generalized alveolar T cell enrichment is associated with survival in severe COVID-19 (*49, 50*), with CD8^+^CD127^HI^ T cells specifically being inversely correlated with SOFA score and duration of mechanical ventilation (*50*). It is tempting to speculate that given the higher proportions of γδ and CD8^+^CD127^HI^ T cells in A-SP2 vs. A-SP4, along with the lower rates of active pneumonia in A-SP2, that individuals in this subphenotype may have mounted more effective host protection against infection, although this hypothesis requires confirmation in mechanistic studies. Together, our findings point toward a very complex interplay of diverse immune cell subsets and mediators within the alveolar space during acute respiratory failure and suggest more refined classifications of these patients are likely necessary to identify effective host-modulating therapies.

There are several knowledge gaps in current models of acute respiratory failure and ARDS subphenotypes – particularly those based on clinical and peripheral blood molecular signatures – that our study helps clarify. The first is that we have confirmed in a large cohort that peripheral blood molecular signatures do not fully capture respiratory biology in critically ill patients (*19, 20, 23–25*). For example, the “hypoinflammatory” ARDS subphenotype, which is consistently identified in approximately two-thirds of patients with ARDS and is associated with better survival compared with the “hyperinflammatory” subphenotype (*18, 53–55*), did not align with alveolar inflammation in many individuals. In our analysis, half of the patients classified as “hypoinflammatory” based on peripheral blood measurements were classified in A-SP4 – the alveolar subphenotype characterized by the highest levels of innate immune cytokines and chemokines (Figure S7). Our data also suggests that the “hypoinflammatory” ARDS subphenotype, A-SP3, and immune exhaustion, either localized to the lung or systemic, do not fully correspond. These findings support the concept that compartmentalization of immune responses is highly relevant in critical illness (*56*), and that discordant responses between the lung and peripheral blood should be considered in the design of immunomodulatory therapies for acute respiratory failure.

A second knowledge gap our study helps clarify is that critical illness subphenotypes are likely very dependent on timing and phase of illness. Our longitudinal analysis indicates that patient assignment into an A-SP is dynamic over the course of a hospitalization (Figure 6). This finding aligns with clinical experience, where the condition of patients with acute respiratory failure often changes hour-by-hour. The UW-ARF Derivation Cohort is a large, real-world clinical cohort where patients were sampled at various times following intubation. To help determine if sampling time was the primary driver of A-SP classification, we stratified patients by intubation-to-BAL interval (Table S3) and sought to validate our findings in two external cohorts composed exclusively of patients with early acute respiratory failure (Table 2). The associations between A-SP assignment and subsequent 28-day mortality may suggest that some alveolar immune signatures are more adaptive than others. However, it is equally plausible that alignment between an alveolar immune profile and a patient’s clinical context may be more important than the specific immune profile itself. Future studies that include larger numbers of patients with repeated samples will be essential to define the alveolar host-response patterns and trajectories most strongly associated with improved outcomes in acute respiratory failure.

Our study has limitations. First, the validation cohorts lacked sufficient power to detect statistically significant mortality differences comparable to those observed in the Derivation Cohort, where a sample size of n = 466 yielded an association that only narrowly met an alpha level of 0.05. Although the overall biologic and clinical profiles of A-SPs were consistent across Derivation and Validation Cohorts – suggesting a robust underlying signal – we recognize that additional validation is essential to establish the generalizability of these subphenotypes. Second, it is not entirely clear whether A-SP1 and A-SP3 represent truly distinct subphenotypes or reflect a “salsa effect,” in which a single population is artificially divided into multiple latent classes (*27*). We acknowledge this possibility; however, several lines of evidence argue that A-SP1 and A-SP3 are biologically and clinically distinct. The proportions of alveolar neutrophils (Figure 2), CXCR3^HI^ T cells, and CD8^+^CD127^HI^ T cells differed significantly between the two groups (Figure 4). We speculate these biologic differences may have contributed to the higher mortality observed in A-SP3 compared with A-SP1 across all three cohorts. Third, BALF sampling may lead to dilutional effects that impact the soluble protein measurements. It is possible that A-SP3, the subphenotype with globally reduced soluble protein levels, may represent individuals with a more dilute BALF sample. The BALF procedure for all three cohorts was protocolized with standardized amounts of saline instilled, which limits this possibility. In addition, the corresponding clinical and alveolar cell profiles strongly suggest subjects in A-SP3 had a less inflammatory alveolar microenvironment than the other A-SPs. A-SP3 had a dramatically lower proportion of alveolar neutrophils compared with the other subphenotypes, which would not be impacted by dilutional factors (Table 2). The clinical severity of respiratory failure as measured by pneumonia, ARDS, S/F ratio, and oxygenation index was also significantly lower in A-SP3 compared with the other A-SPs, again suggesting the corresponding soluble protein profile was not the result of a dilution effect (Figure 2). Fourth, our comparison between A-SPs and the hyper-/hypoinflammatory ARDS subphenotypes was limited to Validation Cohort One, which had available paired BAL and plasma samples. As such, we are unable to fully extrapolate our cellular immunophenotyping findings to existing ARDS subphenotype classifications. Finally, although our soluble protein and spectral flow cytometry panels were large and broadly representative, they were not exhaustive. It is likely additional acute respiratory failure subphenotypes will emerge as more comprehensive molecular measurements – sampled across organs and phases of illness – become available.

In summary, we conducted comprehensive immunophenotyping of lower respiratory specimens and identified novel subphenotypes of acute respiratory failure in one of the largest cohorts of mechanically ventilated patients reported to date. These subphenotypes exhibit distinct biology and associations with clinical outcomes. Over the past decade, a central goal in critical care research has been to define diseases and identify therapeutic targets based on biologic pathways rather than solely on physiology and clinical characteristics. Our findings underscore the importance of incorporating lung-specific biologic measurements into future observational and interventional studies of patients with acute respiratory failure.

## MATERIALS AND METHODS

### Study Design and Participants

This study is an analysis of clinical data and biospecimens collected from subjects representing three cohorts. The UW-ARF Cohort (Derivation Cohort) prospectively enrolled mechanically ventilated subjects at risk for ARDS from 2015 - 2023 who underwent a clinically indicated BAL at Harborview Medical Center (UW Hospital System). Details on the bronchoscopy procedure and sample handling for subjects enrolled in the Derivation Cohort have been published (*22*). The clinical BAL procedure at Harborview Medical Center was standardized during the study time period and consisted of one 20 ml saline lavage waste followed by 4 x 30 ml saline lavages. Research samples were obtained from excess BALF not needed for clinical care under a waiver of informed consent. The Omega-3 Fatty Acid Cohort (Validation Cohort One) was a Phase II randomized controlled trial conducted from 2006 – 2008 that compared omega-3 fatty acid treatment vs. placebo in subjects with early (< 48 hours) ARDS at five North American Centers (*36*). Protocolized research bronchoscopies were performed within 48 hours of ARDS onset, and each subject had paired plasma simultaneously collected with the BALF. Details on the bronchoscopy procedure and sample handling have been published (*36*). The BALI Cohort (Validation Cohort Two) prospectively enrolled mechanically ventilated subjects from 2022 - 2025 who underwent a clinically indicated bronchoscopy with BAL at two hospitals in the Netherlands. We analyzed a subset of patients enrolled into BALI who underwent a clinical bronchoscopy within 48 hours of the start of mechanical ventilation. The UW and Amsterdam UMC Institutional Review Boards approved all studies.

### Soluble Protein Measurements

Information on the assays, dilution factors, quality control metrics, and absolute concentrations for BALF soluble proteins measurements in the Derivation Cohort are shown in Table S1 and Figure S1. BALF soluble protein concentrations were measured using electrochemiluminescent immunoassays per the manufacturer’s instructions (Meso Scale Discovery). Our soluble protein panel was based on prior knowledge and contained a broad range of pyrogens, chemokines, growth factors, cytokines, antibacterial mediators, tolerance, and injury biomarkers. Twenty-five of the original forty proteins met quality control standards and were carried forward for analysis. Information on the soluble protein measurements in Validation Cohorts One and Two have been previously published (*21, 36, 57*). Briefly, IL-6, IL-8 and CSF3 (G-CSF) concentrations were measured with a Luminex bead-based immunoassay (R&D Systems) and sPD-L1 was measured using an MSD assay in Validation Cohort One. In Validation Cohort Two, IL-6, sPD-L1, CSF3 (G-CSF), and VEGF were measured using a Luminex bead-based assay (R&D Systems). We did not normalize individual protein values to a reference measurement to account for dilution due to concern for obscuring biologically relevant variation, which is in accordance with American Thoracic Society Consensus Workshop recommendations (*58*).

### Cellular Immunophenotyping

We analyzed all extant BALF cell samples with > 150,000 leukocytes that had been collected in the UW-ARF Study between 1/1/2016 and 8/1/21 (n = 48) with spectral flow cytometry. BALF supernatant was processed and analyzed for all 466 subjects enrolled in the UW-ARF, however the subset of 48 BALF samples with cryopreserved cells was determined by staff and resource availability at the time of sample acquisition. Details on our cytometric protocols have been published (*22*). Briefly, we aliquoted cells at a concentration of 10^7^ cells/mL freezing media (7% DMSO/FBS). We thawed cryopreserved cells in batches on the day of analysis in a 37°C water bath x 2 minutes, washed, and resuspended cell pellets in Benzonase (1:5000 dilution in RPMI). We stained cells with an eFluor455UV fixable viability dye (Fisher Scientific, Catalogue number: 65-0868-14) x 30 min at room temperature in the dark, washed, and stained the cells with an antibody cocktail x 30min at room temperature. We acquired the cytometric data on a 5-laser Cytek Aurora (Cytek Biosciences) spectral flow cytometer at an event rate for 2500–5000 events per second. We unmixed the spectral flow cytometry data using SpectraFlow software and analyzed the .fcs files with FlowJo version 10. Flow cytometry gating was performed prior to alveolar subphenotype definition assignment.

### Clinical Definitions

Demographics, clinical data, and outcome data in the Derivation Cohort were extracted from the electronic medical record via the University of Washington Data Warehouse. Clinical data in the Validation Cohorts were obtained via their respective case report forms. Clinical BAL cell counts and differentials were assessed through cytospin stains performed in a Clinical Laboratory Improvement Amendment Certified Lab at Harborview Medical Center. These cytospins were performed on fresh samples prior to any cryopreservation. APACHE and SOFA scores were calculated based on their original instruments (*59, 60*). AKI was defined as a ≥ 0.3 mg/dl or 50% from a baseline serum creatinine consistent with KDIGO guidelines (*61*). Shock was defined as receipt of vasopressors on a given calendar day (e.g. bronchoscopy day). ARDS was defined by the 2012 Berlin definition using chest radiographs independently adjudicated by three physicians (NAS, FLM, EDM) (*2*). Oxygenation index (OI) was calculated by dividing the product of F_i_O_2_ and mean airway pressure by the P_a_O_2_ or the S_a_O_2_ (*62*). VFDs were calculated as the number of days a participant was alive and free of invasive mechanical ventilation within the 28-day period after the participant underwent bronchoscopy.

### Statistical Analyses

We excluded alveolar mediators that had > 10% of values outside of the assay’s validated range of detection (Table S1). Analytes measured below the range of detection were imputed at a value 50% the lower limit of detection. All variables were log-transformed and standardized prior to analyses. We considered a final panel of 25 alveolar protein levels obtained in the Derivation Cohort as class-defining variables in the LPA model. We used the VLMR likelihood ratio test, which tests whether class n is a better model than class n-1, as the primary test for model fit. We considered additional criteria, such as BIC (lower values indicating model parsimony) and the entropy statistic (a measure between 0 and 1, with optimal numbers greater than 0.8 indicating good class separation). LPA analysis was performed using Mplus v7.11.

To develop a parsimonious predictive model for A-SP class assignment that we could apply to independent cohorts, we used an extreme gradient boosted model (XGBoost). We performed an unbiased evaluation of the model using 5-fold outer cross-validation, and within each outer training fold we ran an inner cross-validation to tune XGBoost hyperparameters and perform train-only feature ranking. We used the SHAP method to explain the model’s predictions (*63*). We generated SHAP summary plots of the 10 most important predictor variables and ranked them based on their relative contribution to each A-SP assignment. We compared these 4-biomarker parsimonious models against the benchmark of original LPA class assignment in the Derivation Cohort using area under the receiver operating characteristic curves. We carried forward the highest performing available variable for each A-SP to the independent cohorts to assign A-SP.

We used a validated regression-based model with plasma soluble tumor necrosis factor receptor-1 (sTNF-R1), IL-8, and serum bicarbonate concentrations to classify patients in Validation Cohort One into plasma-derived ARDS hyperinflammatory or hypoinflammatory subphenotypes (*23, 54*).

We performed univariable comparisons with T-tests or Mann-Whitney U-tests as appropriate. Categorial clinical variables were compared with Chi-squared tests. We compared the proportion of hyper- and hypoinflammatory ARDS across the alveolar subphenotypes using a Fisher’s exact test given the sample size. The Benjamini-Hochberg (BH) procedure was used to correct for multiple hypothesis testing in univariable analyses when noted. Some adjusted *p*-values per subset comparison were identical due to the monotonicity correction within BH procedure, small number of tests, and range of similar *p*-values. Survival analyses were done by Kaplan-Meier estimation implemented in the survival method (R packages: *survival_3.8-3* and *survminer_0.5.0*). We estimated relative risk (RR) of 28-day mortality using Poisson regression with robust standard errors adjusting for age, sex, pneumonia, ARDS, and interval between intubation and BAL day using the *glm* (generalized linear model) function in R. A *p*-value or *p adjusted*-value < 0.05 was the threshold for statistical significance. We generated datasets using Python3, created figures in GraphPad Prism version 10, and performed all statistical analyses in R version 4.5.0.

## Data Availability

All data produced in the present study are available upon reasonable request to the authors

## Acknowledgements

The authors thank the research participants, their families, and the staff at HMC for their generous participation in our study. We want to acknowledge the essential role Jillian Simmons played in obtaining BAL samples. We would also like to acknowledge the BRI Cell and Tissue Analysis Core (RRID: SCR_026327) for flow cytometry support including Dr. Adam Wojno (Director). We also thank the M.J. Murdock Charitable Trust for generously providing equipment funding for the BRI Cores. Finally, we thank the late Dr. Thomas Martin who directed the UW P50 SCORE Grant that helped support the Omega-3 Fatty Acid Trial and for his long-term mentorship.

## Funding

National Institutes of Health grant NHLBI R01 HL149676 (CM, MMW)

National Institutes of Health grant NHLBI K23HL144916 (EDM)

National Institutes of Health grant NHLBI R01 HL169265 (EDM, AK, CLH, MMW, CM)

National Institutes of Health grant NHLBI P50 HL073997 (RDS, MMW)

National Institutes of Health grant NHLBI R01 HL172872 (PKB)

National Institutes of Health grant NHLBI K23 HL 175120 (SEH)

Parker B. Francis Foundation (SEH)

European Respiratory Society Short-Term Research Fellowship STRTF202310-01115 (LSB)

## Author Contributions

*Study concept and design:* EDM, CM

*Protocol development and acquisition, analysis, and interpretation of data:* EDM, TL, MO, LSB, NAS, CS, MM, AC, LYG, FLM, SEH, PKB, AK, CLH, RDS, HV, JWD, LDB, MMW, CM

*Mediator measurements and analysis:* CS, MM, AC, RDS, MMW, HV, LSB, JWD, LDB, EDM, CM

*Flow cytometry measurements and analysis*: MO, EDM, CM

*Computational and statistical analysis:* TL, NAS, EDM

*Clinical adjudication and analysis*: NAS, FLM, EDM, CM, RDS, MMW, HV, LSB, LDB

*Literature search and initial drafting of the manuscript:* EDM, CM

*Contributed to and approve the final version of the manuscript:* EDM, TL, MO, LSB, NAS, CS, MM, AC, LYG, FLM, SEH, PKB, AK, CLH, RDS, HV, JWD, LDB, MMW, CM

*Agree to be accountable for all aspects of the work:* EDM, TL, MO, LSB, NAS, CS, MM, AC, LYG, FLM, SEH, PKB, AK, CLH, RDS, HV, JWD, LDB, MMW, CM

## Competing Interests

The authors declare no competing interests.

## Data and Materials Availability

All data are available in the article and its Supplementary files or from the corresponding author upon request.

## Artificial Intelligence Statement

ChatGPT-5 was used for minor editing of final versions of manuscript drafts to help conform to word count limits. No artificial intelligence tools were used for literature search, analysis, data interpretation, data presentation, or primary drafting of the manuscript.

## LIST OF SUPPLEMENTARY MATERIALS

**Figure S1.**
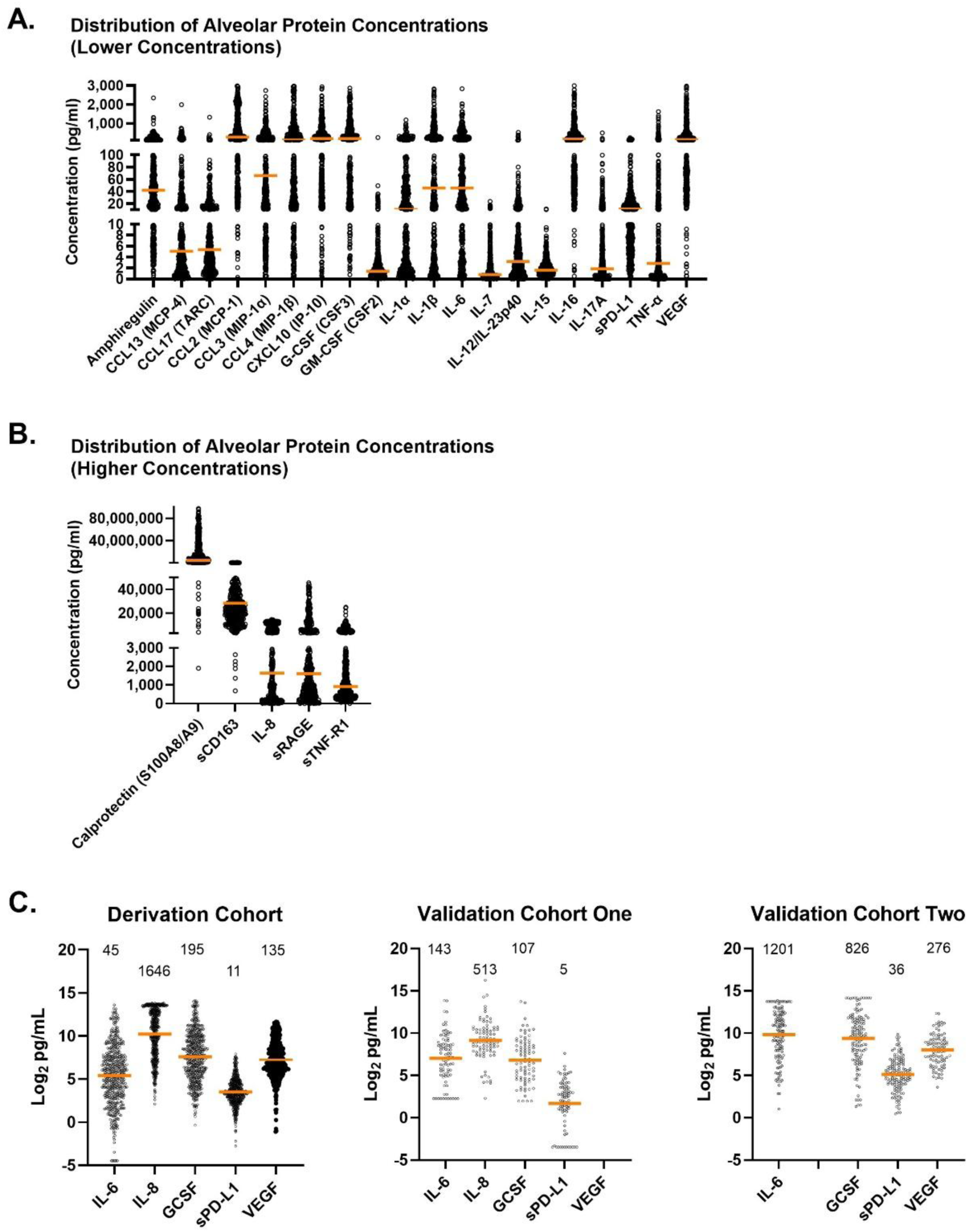
Concentrations of Alveolar Proteins. BALF concentrations of respective proteins in the Derivation Cohort and Validation Cohorts One and Two. Displayed are the individual values, median, and interquartile range for each soluble protein. **A)** Derivation Cohort – proteins with low/moderate BALF concentrations; **B)** Derivation Cohort – proteins with high BALF concentrations; **C)** Log_2_-transformed concentrations of the select proteins used in the 4-biomarker parsimonious models across cohorts. The median non-transformed concentration of each protein is displayed above the individual values.

**Figure S2.**
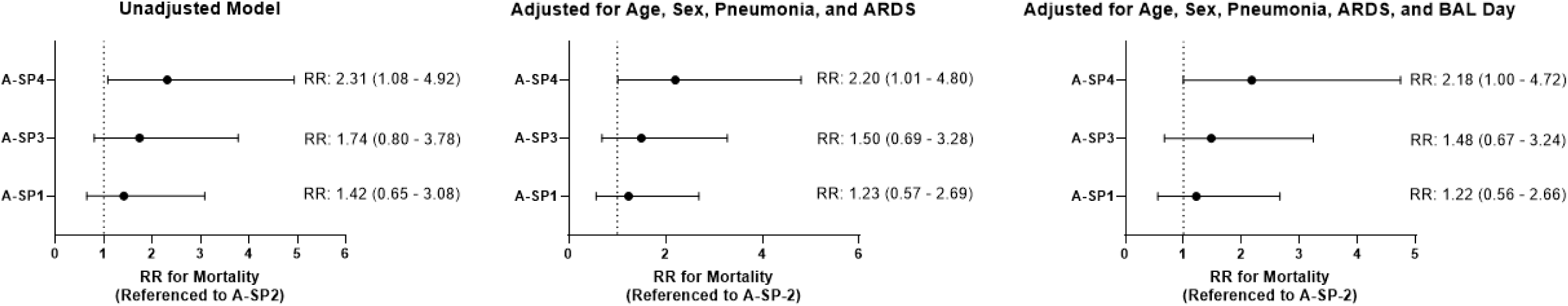
Adjusted Models Testing for Associations Between Alveolar Subphenotypes and Mortality. Forrest plots display the relative risk (RR) and 95% confidence interval for mortality by 28 days following BAL in patients with acute respiratory failure. Each A-SP displayed on the Y-axis is compared with A-SP2 (the reference).

**Figure S3.**
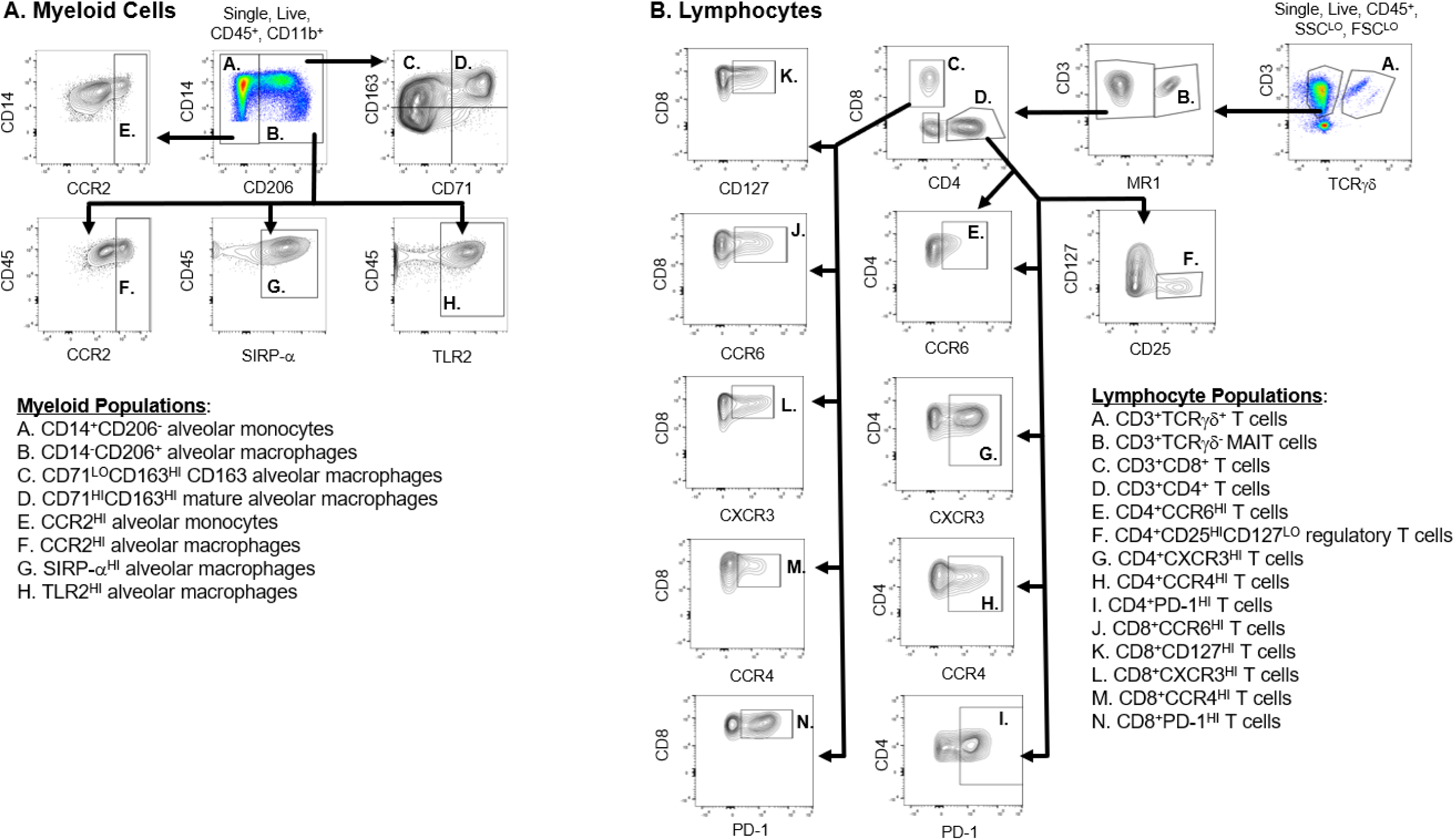
Alveolar Leukocyte Subset Gating. We used spectral flow cytometry to classify alveolar leukocytes collected from subjects with acute respiratory failure. Representative gating of: **A)** single, live, CD45^+^, CD11b^+^ alveolar myeloid cells; or **B)** single, live, CD45^+^, low forward and side scatter alveolar lymphocytes

**Figure S4.**
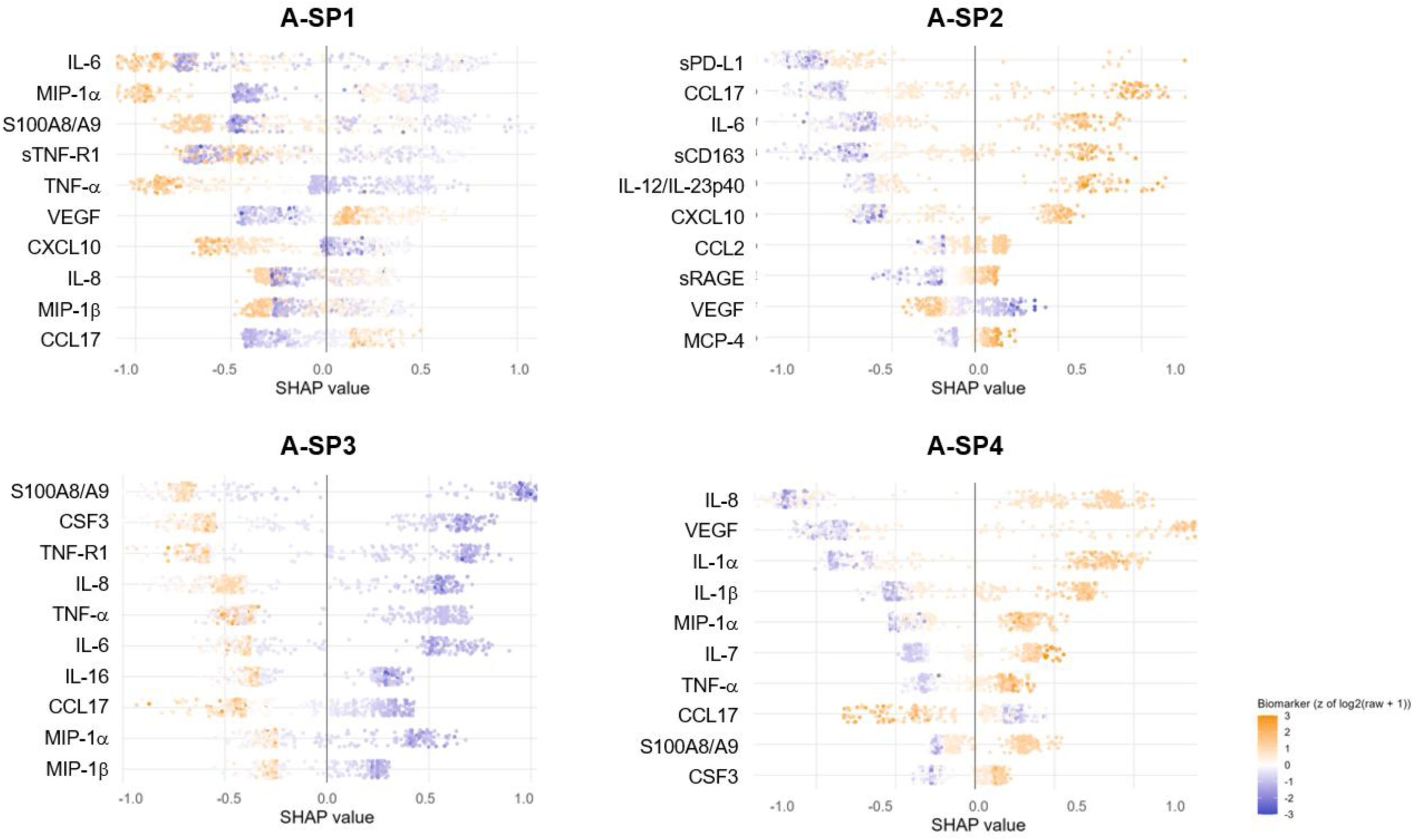
SHAP Summary Plot of the 10 Most Important Predictors for Each Alveolar Subphenotype. Shapley additive explanation (SHAP) values for each A-SP are plotted on the x-axis. Each dot represents an individual observation. The value of each predictor is represented by the colors. Orange indicates high levels of each mediator and blue represents low levels. The importance of the predictors to the overall model is determined by the predictors’ absolute total SHAP value across all patients and is displayed in descending rank-order from high-to-low on the Y-axis.

**Figure S5.**
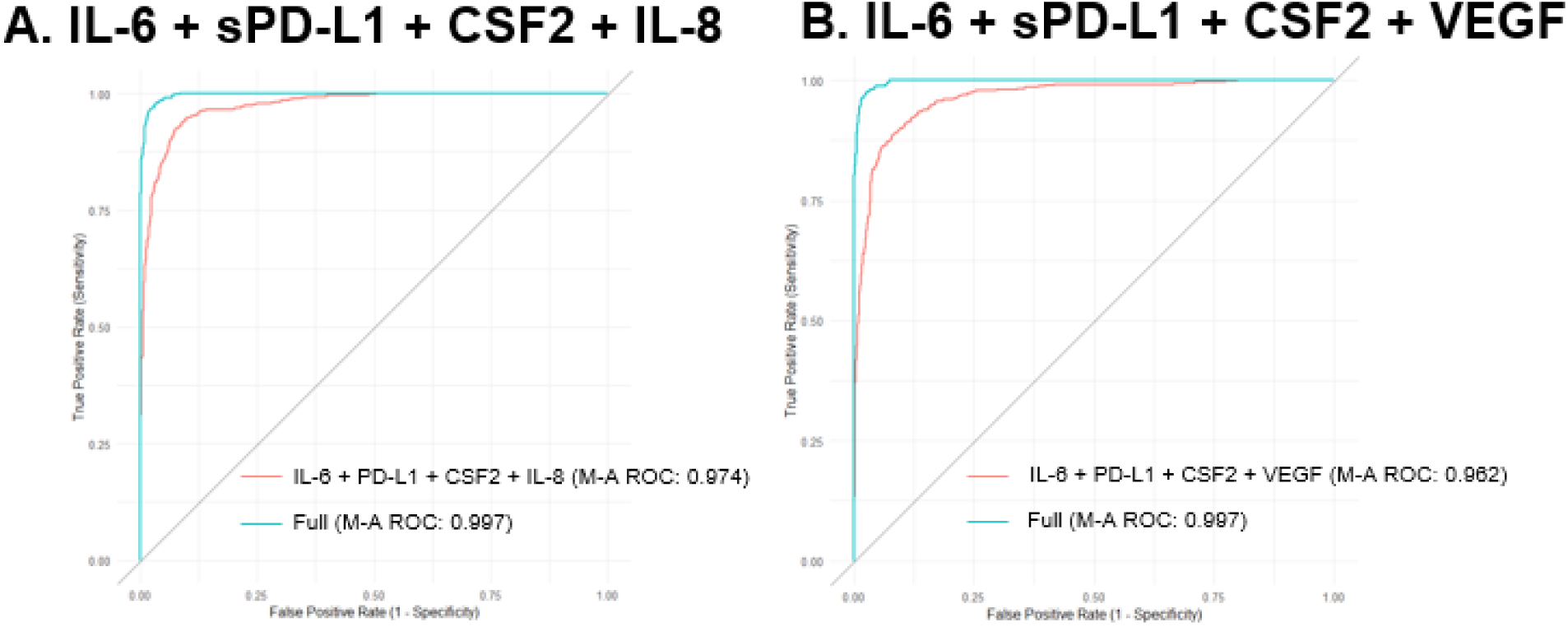
The Performance of Four-Biomarker Models in Derivation Cohort. Receiver operating curves for parsimonious alveolar subphenotype (A-SP) prediction models. Area under the receiver operating characteristic curve (AUC) for A-SP class assignment using two different 4-biomarker models (**panels A and B**) compared with a full model using 25 alveolar mediators in the Derivation Cohort. Each model is benchmarked against the gold standard of original A-SP classification using latent profile analysis.

**Figure S6.**
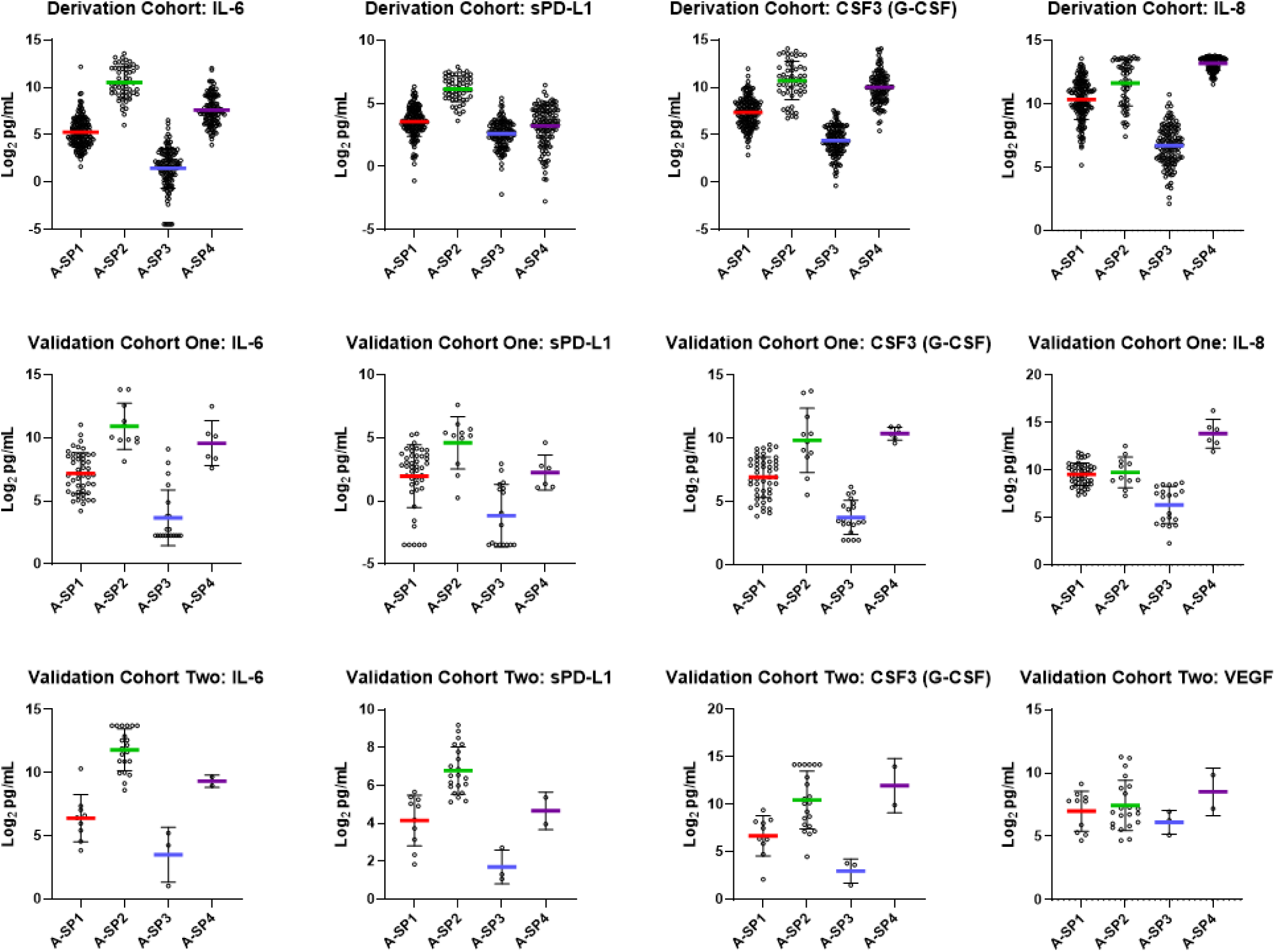
Alveolar Protein Concentrations Between A-SPs and Cohorts. Log_2_ transformed BALF concentrations of respective proteins in the Derivation Cohort and Validation Cohorts One and Two. Displayed are the individual values, mean, and standard deviation for each protein between each alveolar subphenotype (A-SP)

**Figure S7.**
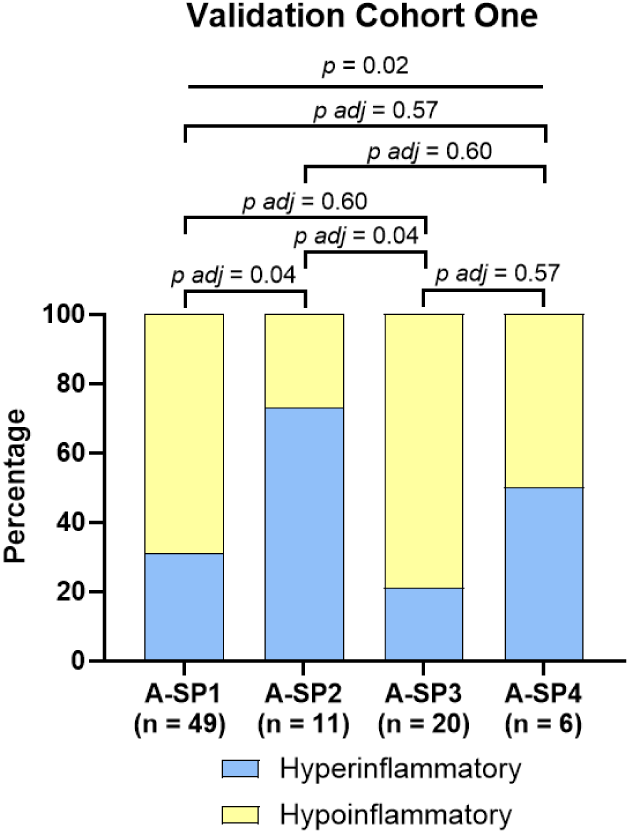
Overlap Between Alveolar and ARDS Subphenotypes. Bar plot comparing the proportion of patients with “hyperinflammatory” ARDS between A-SPs using a Fisher’s test with post-hoc pairwise proportion tests with FDR correction.

**Table S1.** Protein Measurement Assay Performance.

| Analyte | Catalog Number | Dilution Factor | Intraplate % CV | Interplate % CV | Below LLOD (%) | Passed QC† |
| --- | --- | --- | --- | --- | --- | --- |
| Amphiregulin | F21AAE-3 | Neat | 5.52 | 6.29 | 0.00 | Yes |
| Calprotectin (S100A8/A9) | K151AJYR-2 | 1:1000 | 9.87 | 23.73 | 0.20 | Yes |
| CCL13 (MCP-4) | K15047G | 1:2 | 6.29 | 12.16 | 0.20 | Yes |
| CCL17 (TARC) | K15047G | 1:2 | 14.62 | 47.09 | 0.60 | Yes* |
| CCL2 (MCP-1) | K15047G | 1:2 | 4.74 | 11.34 | 0.40 | Yes |
| CCL3 (MIP-1α) | K15047G | 1:2 | 5.18 | 11.02 | 1.59 | Yes |
| CCL4 (MIP-1β) | K15047G | 1:2 | 13.02 | 19.99 | 0.40 | Yes |
| sCD163 | K151J4R-2 | 1:8 | 6.39 | 15.11 | 0.20 | Yes |
| CXCL10 (IP-10) | K15047G | 1:2 | 3.78 | 13.25 | 0.00 | Yes |
| G-CSF (CSF3) | K151XXK | Neat | 8.27 | 9.67 | 0.20 | Yes |
| GM-CSF (CSF2) | K15050G | 1:2 | 4.89 | 15.86 | 0.00 | Yes |
| IL-1α | K15050G | 1:2 | 5.88 | 19.99 | 0.00 | Yes |
| IL-1β | K15049G | 1:8 | 4.77 | 8.47 | 0.60 | Yes |
| IL-6 | K15049G | 1:8 | 5.04 | 15.66 | 1.20 | Yes |
| IL-7 | K15050G | 1:2 | 6.49 | 12.06 | 0.60 | Yes |
| IL-8 | K15049G | 1:8 | 3.64 | 12.95 | 0.00 | Yes |
| IL-12/IL-23p40 | K15050G | 1:2 | 3.56 | 5.18 | 1.59 | Yes |
| IL-15 | K15050G | 1:2 | 7.93 | 12.32 | 0.20 | Yes |
| IL-16 | K15050G | 1:2 | 3.35 | 9.87 | 0.00 | Yes |
| IL-17A | K15050G | 1:2 | 3.59 | 9.47 | 6.94 | Yes |
| sPD-L1 | K151Z7K | 1:2 | 2.71 | 5.87 | 0.00 | Yes |
| sRAGE | K151AHAK-2 | 1:18 | 4.08 | 10.63 | 0.00 | Yes |
| TNF-R1 | K151AHQK | 1:8 | 2.13 | 7.47 | 0.00 | Yes |
| TNF-α | K15049G | 1:8 | 6.31 | 8.35 | 7.77 | Yes |
| VEGF | K15050G | 1:2 | 3.50 | 4.27 | 0.00 | Yes |
| Ang-1 | K15227N-2 | 1:2 | 4.19 | 4.19 | 84.91 | No |
| Ang-2 | K15227N-2 | 1:2 | 10.56 | 17.10 | 37.61 | No |
| ANGPTL-4 | K151Z3R | 1:2 | 2.76 | 11.08 | 16.87 | No |
| CCL11 (Eotaxin) | K15047G | 1:2 | 38.52 | 60.51 | 11.73 | No |
| CCL22 (MDC) | K15047G | 1:2 | 29.38 | 47.03 | 4.97 | No |
| CCL26 (Eotaxin-3) | K15047G | 1:2 | 47.82 | 362.79 | 15.71 | No |
| IFN-γ | K15049G | 1:8 | 42.75 | 253.72 | 43.03 | No |
| IL-10 | K15049G | 1:8 | 7.54 | 9.30 | 13.75 | No |
| IL-12p70 | K15049G | 1:8 | 60.56 | 87.57 | 26.49 | No |
| IL-13 | K15049G | 1:8 | 15.22 | 31.02 | 11.75 | No |
| IL-2 | K15049G | 1:8 | 29.96 | 111.56 | 11.55 | No |
| IL-3 | K151WZD | Neat | 8.95 | 66.10 | 36.49 | No |
| IL-4 | K15049G | 1:8 | 36.18 | 74.83 | 43.23 | No |
| IL-5 | K15050G | 1:2 | 13.34 | 24.57 | 7.54 | No |
| TNF-β | K15050G | 1:2 | 29.13 | 87.58 | 37.50 | No |

CV = coefficient of variation; LLOD = lower limit of detection
† Criteria: 1) > 10% of samples below LLOD; 2) intraplate CV > 15%; or 3) interplate CV > 25%
\* During our QC assessment, we determined that two of the plates measuring CCL17 (TARC) had bubbles in control wells that led to high interplate %CV. Based on this assay's prior performance, we elected to include this analyte in our subsequent analyses.

**Table S2.** Optimal Alveolar Biomarker Classes.

| Classes | BIC | Entropy | VLMR<br>p-value | Number of Individuals Per Class |  |  |  |  |
| --- | --- | --- | --- | --- | --- | --- | --- | --- |
|  |  |  |  | N1 | N2 | N3 | N4 | N5 |
| 2 | 28368.221 | 0.973 | 0 | 219 | 247 |  |  |  |
| 3 | 26555.993 | 0.958 | 0.0397 | 170 | 133 | 163 |  |  |
| **4** | **25151.848** | **0.969** | **0.0182** | **161** | **55** | **131** | **119** |  |
| 5 | 25038.763 | 0.97 | 0.0557 | 114 | 107 | 54 | 1051 | 86 |

BIC = Bayesian Information Criteria; VLMR = Vuong-Lo-Mendell-Rubin test

**Table S3.** Sensitivity Analyses of Associations Between Alveolar Subphenotypes (A-SPs) and Mortality.

|  | **A-SP1**<br>**(n = 161)** | **A-SP2**<br>**(n = 55)** | **A-SP3**<br>**(n = 131)** | **A-SP4**<br>**(n = 119)** | **Total**<br>**(n = 466)** |
| --- | --- | --- | --- | --- | --- |
| **Mortality in Each Alveolar Subphenotype Stratified by Intubation-to-BAL Interval** |  |  |  |  |  |
| BAL Day<br>0 – 6 | 14/90<br>**(15.5%)** | 6/43<br>**(14.0%)** | 11/58<br>**(19.0%)** | 16/65<br>**(24.6%)** | 47/256<br>**(18.4%)** |
| BAL Day<br>7 – 14 | 8/41<br>**(19.5%)** | 1/11<br>**(9.1%)** | 12/38<br>**(31.6%)** | 11/33<br>**(33.3%)** | 32/123<br>**(26.0%)** |
| BAL Day<br>> 14 Days | 7/30<br>**(23.3%)** | 0/1<br>**(0%)** | 6/35<br>**(17.1%)** | 8/21<br>**(38.1%)** | 21/87<br>**(24.1%)** |
| **Mortality in Each Alveolar Subphenotype Stratified by Median Age** |  |  |  |  |  |
| < 51.52 Years | 7/70<br>**(10%)** | 2/38<br>**(5.3%)** | 7/60<br>**(11.7%)** | 16/65<br>**(24.6%)** | 32/233<br>**(13.7%)** |
| > 51.52 Years | 22/91<br>**(24.2%)** | 5/17<br>**(29.4%)** | 22/71<br>**(31.0%)** | 19/54<br>**(35.2%)** | 68/233<br>**(29.2%)** |
| **Mortality in Each Alveolar Subphenotype Stratified by Pneumonia Status** |  |  |  |  |  |
| No<br>Pneumonia | 16/65<br>**(24.6%)** | 4/29<br>**(13.8%)** | 16/81<br>**(19.8%)** | 7/15<br>**(46.7%)** | 43/190<br>**(22.6%)** |
| Yes<br>Pneumonia | 13/96<br>**(13.5%)** | 3/26<br>**(11.5%)** | 13/50<br>**(26.0%)** | 28/104<br>**(26.9%)** | 57/276<br>**(20.7%)** |
| **Mortality in Each Alveolar Subphenotype Stratified by ARDS** |  |  |  |  |  |
| No<br>ARDS | 10/71<br>**(14.1%)** | 4/15<br>**(26.7%)** | 14/73<br>**(19.2%)** | 11/48<br>**(22.9%)** | 39/207<br>**(18.8%)** |
| Yes<br>ARDS | 19/90<br>**(21.1%)** | 3/40<br>**(7.5%)** | 15/58<br>**(25.9%)** | 24/71<br>**(33.8%)** | 61/259<br>**(23.6%)** |
| **Mortality in Each Alveolar Subphenotype Stratified by Corticosteroids** |  |  |  |  |  |
| No<br>Steroids | 22/140<br>**(15.7%)** | 6/43<br>**(14.0%)** | 26/115<br>**(22.6%)** | 29/110<br>**(26.4%)** | 83/408<br>**(20.3%)** |
| Yes<br>Steroids | 7/21<br>**(33.3%)** | 1/12<br>**(8.3%)** | 3/16<br>**(18.8%)** | 6/9<br>**(66.7%)** | 17/58<br>**(29.3%)** |
| **Mortality in Each Alveolar Subphenotype Stratified by Antibiotics** |  |  |  |  |  |
| No<br>Antibiotics | 7/72<br>**(9.7%)** | 4/20<br>**(20.0%)** | 14/61<br>**(23.0%)** | 20/54<br>**(37.0%)** | 45/207<br>**(21.7%)** |
| Yes<br>Antibiotics | 22/89<br>**(24.7%)** | 3/35<br>**(8.6%)** | 15/70<br>**(21.4%)** | 15/65<br>**(23.1%)** | 55/259<br>**(21.2%)** |
| **Total** |  |  |  |  |  |
| **Total** | 29/161<br>**(18.0%)** | 7/55<br>**(12.7%)** | 29/131<br>**(22.1%)** | 35/119<br>**(29.4%)** | 100/466<br>**(21.5%)** |

Mortality censored at 28 days following BAL; pneumonia was defined in BALs with a pathogen growing $\geq 10,000$ cfu/ml; corticosteroids defined as receipt of dexamethasone, methylprednisolone, hydrocortisone, or prednisone on bronchoscopy day or day prior to bronchoscopy day; antibiotics defined as receipt of any antibiotics within 48 hours prior to bronchoscopy
A-SP = alveolar subphenotype; BAL = bronchoalveolar lavage; BAL Day = number of days between intubation and the BAL

**Table S4.**
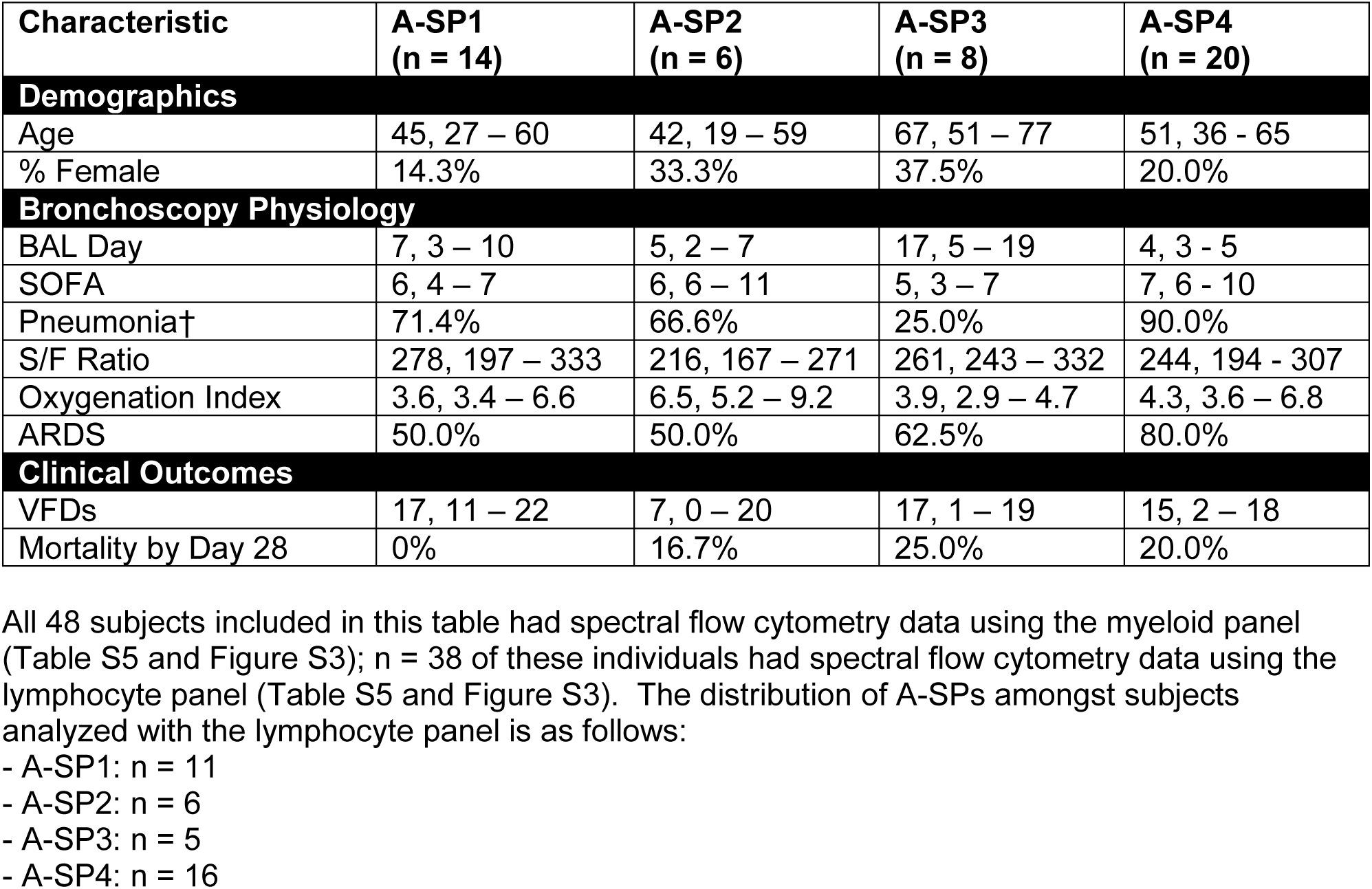
Clinical Characteristics of Subjects with Spectral Flow Cytometry Data.

**Table S5.** Spectral Flow Cytometry Cell Surface Protein Panel.

| Cell Surface Protein | Clone | Conjugated Fluorophore | Manufacturer | Catalog Number |
| --- | --- | --- | --- | --- |
| **Myeloid Panel** |  |  |  |  |
| CD45 | 2D1 | AF700 | Biolegend | 368513 |
| CD3 | UCHT1 | BV570 | Biolegend | 300436 |
| CD11b | ICRF44 | APC | Biolegend | 301309 |
| CD14 | M5E2 | APC/Cyanine7 | Biolegend | 301819 |
| CD15 | HI98 | BUV395 | BD Biosciences | 563872 |
| CD71 | CY1G4 | BV650 | Biolegend | 334115 |
| CD163 | GHI/61 | BV605 | Biolegend | 333616 |
| CD172a (SIRP-a) | 15-414 | FITC | Biolegend | 372107 |
| CD192 (CCR2) | K036C2 | PE/Dazzle 594 | Biolegend | 357221 |
| CD169 | 7-239 | BV421 | Biolegend | 346017 |
| CD206 | 15-2 | BV785 | Biolegend | 321142 |
| CD274 (PD-L1) | 29E.2A3 | BV711 | Biolegend | 329722 |
| CD282 (TLR2) | TL2.1 | PE | Biolegend | 309707 |
| CD326 (EpCAM) | 9C4 | PE/Cyanine7 | Biolegend | 324221 |
| **Lymphocyte Panel** |  |  |  |  |
| gdTCR | B1 | BV421 | Biolegend | 331218 |
| CD8 | SK1 | V500 | BD Biosciences | 561618 |
| CD45 | HI30 | BV570 | Biolegend | 304033 |
| CD279 (PD-1) | A17188B | APC | Biolegend | 621610 |
| CD196 (CCR6) | 11A9 | APC-R700 | BD Biosciences | 565173 |
| CD25 | M-A251 | APC-Fire810 | Biolegend | 356150 |
| CD183 (CXCR3) | G025H7 | PE-Cy7 | Biolegend | 353720 |
| MR1 | 5-OP-RU | PE | NIH Tetramer Facility | 54466 (5-OP-RU) |
| CD4 | SK3 | cFluorYG584 | Cytek | R7-20042 |
| CD127 | A019D5 | AF488 | Biolegend | 351313 |
| CD194 (CCR2) | 1G1 | CD786 | BD Biosciences | 744141 |
| CD45RA | HI100 | BUV805 | BD Biosciences | 742020 |
| HLA-DR | L243 | BV650 | Biolegend | 307650 |

**Table S6.** Clinical Characteristics of Validation Cohorts One and Two.

| Characteristic | Total<br>(n = 86) | Total<br>(n = 36) |
| --- | --- | --- |
| **Demographics** |  |  |
| Age | 50, 41 – 63 | 56, 47 - 66 |
| % Female | 37% | 39% |
| **Peri-Bronchoscopy Physiology** |  |  |
| BAL Day | < 48 hours | < 48 hours |
| Omega-3 FA | 44.2% | NA |
| P/F Ratio | 163, 126 - 205 | 151, 100 - 189 |
| ARDS | 100% | 89% |
| **Clinical Outcomes** |  |  |
| VFDs | 14, 0 – 21 | 0, 0 - 23 |
| 14-Day Mortality | 9.3% | 27.8% |
| 28-Day Mortality | 16.3% | 41.7% |

ARDS = acute respiratory distress syndrome; BAL = bronchoalveolar lavage; Omega-3 FA (fatty acid) = percentage of patients randomized to the treatment group; P/F ratio = $P_aO_2/F_iO_2$ ratio; VFDs = ventilator-free days

## Notes

### Competing Interest Statement

The authors have declared no competing interest.

### Author Declarations

The University of Washington Human Subjects Division and the Amsterdam UMC IRB gave ethical approval for this work.

